## Supplementary Figs S1-S34, Supplementary Tables S1-S3 for "Maternal pertussis immunization and the blunting of routine vaccine effectiveness: A meta-analysis and modeling study"

#### Table of contents

**S1 Supplementary Materials and Methods**
1.1 Model schematic
1.2 Model equations
1.3 Estimation of relative risk of infection following maternal immunization: mathematical equations
**S2 Supplementary Figures**
Figs. S1-S34
**S3 Supplementary Tables**
Tables S1-S3
**S4 References**

### S1 Supplementary Materials and Methods

#### S1.1 Explanation of model schematic

We implemented an age-structured model of pertussis transmission, extending the previously described model, based on data in Massachusetts [1]. In brief, the model is an extension of the standard SEIR model, in which we follow groups of individuals according to their immunization history, including that of their mothers during pregnancy (Fig. S1). Newborns can be born from immunized mothers ($M$ followed at later ages by superscript$^{(M)}$), mothers whose immunization failed ($S^{(\bar{M})}$or at later ages any compartment with superscript$^{(\bar{M})}$) or from unvaccinated mothers $S$. Maternal immunization occurs at a coverage $p_0(t)$ and with an effectiveness $\varepsilon_M$. Distinguishing between mothers whose immunization failed and mothers who were not immunized is needed when estimating the RR of contracting pertussis because ignoring the failure of maternal immunization results in an overestimation of the effectiveness of maternal immunization and possibly of blunting. This becomes clear in the estimation of RR (see formula in section 1.3), where PCV (proportion of cases vaccinated) is the proportion of cases in infants vaccinated born to vaccinated mothers among all cases in infants vaccinated.

The time step of our model is 1 day. For each time step, individuals remain in one of their 'birth compartments' (unless stated otherwise) until primary immunization. We chose a pertussis immunization schedule that closely resembles that of the empirical studies for which we obtained the RR estimates (Table S1). Although primary immunization often consists of 2 or generally 3 doses with a few months interval, such as 2,3, and 4 months or 2, 4, and 6 months (Table S1), for simplicity, we modeled the infant primary immunization as one event at the age of 3 months. Hence, infants who receive their primary immunization in our model match infants receiving the three doses of primary immunization in a real world setting. In our model, infants receive their primary pertussis immunization (V or superscript$^{(V)}$) at a coverage $p_1(t)$ and with an effectiveness $\varepsilon$. Consistent with the fact that there are three compartments of maternal immunization, there are three compartments for successful primary immunization (Fig. S2: $V^{(M)}$, $V^{(\bar{M})}$and $V$), and three compartments for failed primary immunization, thereby becoming susceptible (Fig. S2: $M^{(V)}$, $S^{(V\bar{M})}$and $S^{(V)}$).

Following primary immunization, we added a pediatric booster at the age of 1.5 years, which closely resembles that of two out of three empirical studies for which a booster is given between the age of 1 and 1.5 years (see Table S1 and references therein). This pediatric booster is given at a probability $p_2(t)$ and with an effectiveness $\varepsilon$. This booster is modeled such that individuals without immunity (because of failed immunization or because of waning) go from their respective aforementioned susceptible compartments to their respective aforementioned immunized compartments.

The schedule of immunization is modeled by moving a fraction of susceptibles to the vaccinated class when individuals reach the following age groups:

- Age 0–2 mo ($i = 1$, maternal immunization): $p_0(t) = v_0\mathbb{I}(t \geq t_M)$, where $v_0$ is the proportion of mothers vaccinated during pregnancy and $t_M$ the start time of maternal immunization.
- Age 3–18 mo ($i = 2$, primary immunization): $p_1(t) = v_1\mathbb{I}(t \geq t_V)$, where $v_1$ is the vaccine coverage for the primary immunization and $t_V$ the start time of vaccination.
- Age 1.5 yr ($i = 3$, pediatric booster): $p_2(t) = v_2\mathbb{I}(t \geq t_V)$, where $v_2$ is the vaccine coverage for booster doses.

After these three age classes, and as in [1, 2], individuals are divided into 1-year age groups from age 2 (i.e., 2 to <3) years to age 74 (i.e., 74 to <75) years. Hence, altogether, the model consists of 76 age groups, labeled $i = 1, \ldots, 76$. Aging occurs continuously, at rates $\delta_i = 1/\text{age span}$, i.e., $\delta_1 = 12/2$ yr$^{-1}$ in newborns, $\delta_2 = 12/15$ yr$^{-1}$ in infants aged 3–18 months, $\delta_3 = 12/6$ yr$^{-1}$ in infants aged between 18 months and 2 years, and $\delta_{i\geq 4} = 1$ yr$^{-1}$ in older age groups.

Following the immunological evidence showing that the presence of maternal antibodies cause an immediate reduction in the antibody response to primary immunization [3, 4, 5], we introduced blunting in the model through a failure of initial vaccine effectiveness: in children immunized in the presence of maternal antibodies, the initial vaccine effectiveness is reduced by a factor $b_1$, resulting in an blunted vaccine effectiveness $\bar{\varepsilon} = \epsilon(1 - b_1)$ (Fig. 1). When infants receive their primary immunization, the antibody concentrations of infants from vaccinated mothers are 30% to 60% lower compared to those from unvaccinated mothers [4, 5]. However, there are no studies available on the extent to which such changes in IgG titers translate into changes in protection. In our study, we chose a decrease in the effectiveness of primary immunization (i.e., protection) from 0% (i.e., no blunting) to 20% (see Fig. 4 and Fig. 5).

In all simulations, we assumed a type-I mortality (whereby all individuals reach age 74 and die at age 75) and no migration. Given these assumptions, the total population size, $N = 10^7$, is held approximately constant by fixing the birth rate $\mu = \frac{1}{75}$ per year. The force of infection in age group $i$ is given by:

$$\lambda_i = q_i \sum_{j=1}^{15} F_{ij}(t) \tilde{C}_{ij} \frac{\tilde{I}_j + \iota}{N_j}$$

where $q_i$ is the probability of infection given exposure (i.e., susceptibility) in age group $i$, $N$ is the total population in age group $j$, and $\iota$ an immigration term, fixed to 1 in all simulations.

The contact matrix $C = (C_{i,j})_{i=1,\ldots,15, j=1,\ldots,15}$ was available from the POLYMOD study in Great Britain, after application of a correction to ensure reciprocity of contacts between age groups. The matrix $C = (C_{i,j})_{i=1,\ldots,76, j=1,\ldots,15}$ represents the full contact matrix, augmented to incorporate the extra age groups in the model, under the assumption that all individuals within a 5-yr age block have similar contact rates. The terms $\tilde{I}_j$ represent the numbers of infected individuals, aggregated over 5-yr age blocks (0–4, 5–9, ..., 70–74) to match the age groups in the POLYMOD study.

The seasonality in children's contact was modeled using the age-dependent seasonal transmission terms $F_{ij}(t)$. Following[1], this forcing was assumed assortative and applied only within the same 5-yr age group, that is, only for contacts between children 5–9 yr and 5–9 yr, between 10–14 yr and 10-14 yr, and between 15–19 yr and 15–19 yr. The seasonality coefficients were fixed according to the estimates obtained in[1], with seasonal maxima of transmission in May for 5–9 year old and September for 10–19 year old (Fig. S8 in[1]).

#### S1.2 Model equations

The deterministic dynamics in newborns are governed by the following system of ordinary differential equations:

$$
\begin{aligned}
\dot{V}_1^{(M)} &= 0 \\
\dot{V}_1^{(\bar{M})} &= 0 \\
\dot{V}_1 &= 0 \\
\dot{M}_1 &= p_0(t)\varepsilon_M \mu N - (\tau + \delta_1)M_1 \\
\dot{M}_1^{(V)} &= 0 \\
\dot{S}_1^{(\bar{M})} &= p_0(t)(1 - \varepsilon_M)\mu N + \tau M_1 - (\lambda_1(t) + \delta_1)S_1^{(\bar{M})} \\
\dot{S}_1 &= (1 - p_0(t))\mu N - (\lambda_1(t) + \delta_1)S_1 \\
\dot{E}_1 &= \lambda_1(t)S_1 - (\sigma + \delta_1)E_1 \\
\dot{I}_1 &= \sigma E_1 - (\gamma + \delta_1)I_1 \\
\dot{S}_1^{(V)} &= 0 \\
\dot{S}_1^{(V\bar{M})} &= 0 \\
\dot{S}_1^{(VM)} &= 0 \\
\dot{R}_1 &= \gamma I_1 - \delta_1 R_1
\end{aligned}
$$

In the second age group ($i = 2$, receipt of primary vaccine course), the equations are:

$$
\begin{aligned}
\dot{V}_2^{(M)} &= \delta_1 V_1^{(M)} + p_1(t)\bar{\varepsilon}\delta_1 M_1 - (\alpha_V + \delta_2)V_2^{(M)} \\
V_2^{(\bar{M})} &= \delta_1 V_1^{(\bar{M})} + p_1(t)\varepsilon\delta_1 S_1^{(\bar{M})} - (\alpha_V + \delta_2)V_2^{(\bar{M})} \\
\dot{V}_2 &= \delta_1 V_1 + p_1(t)\varepsilon\delta_1 S_1 - (\alpha_V + \delta_2)V_2 \\
\dot{M}_2 &= \delta_1 M_1 - p_1(t)\delta_1 M_1 - (\tau + \delta_2)M_2 \\
\dot{M}_2^{(V)} &= \delta_1 M_1^{(V)} + p_1(t)[1 - \bar{\varepsilon}]\delta_1 M_1 - (\tau + \delta_2)M_2^{(V)} \\
\dot{S}_2^{(\bar{M})} &= \delta_1 S_1^{(\bar{M})} - p_1(t)\delta_1 S_1^{(\bar{M})} + \tau M_2 - (\lambda_2(t) + \delta_2)S_2^{(\bar{M})} \\
\dot{S}_2 &= \delta_1 S_1 - p_1(t)\delta_1 S_1 - (\lambda_2(t) + \delta_2)S_2 \\
\dot{E}_2 &= \delta_1 E_1 + \lambda_2(t)(S_2 + S_2^{(\bar{M})} + S_2^{(V)} + S_2^{(V\bar{M})} + S_2^{(VM)}) - (\sigma + \delta_2)E_2 \\
\dot{I}_2 &= \delta_1 I_1 + \sigma E_2 - (\gamma + \delta_2)I_2 \\
\dot{S}_2^{(V)} &= \delta_1 S_1^{(V)} + p_1(t)[1 - \varepsilon]\delta_1 S_1 + \alpha_V V_2 - (\lambda_2(t) + \delta_2)S_2^{(V)} \\
\dot{S}_2^{(V\bar{M})} &= \delta_1 S_1^{(V\bar{M})} + \alpha_V V_2^{(\bar{M})} + p_1(t)[1 - \varepsilon]\delta_1 S_1^{(\bar{M})} - (\lambda_2(t) + \delta_2)S_2^{(V\bar{M})} \\
\dot{S}_2^{(VM)} &= \delta_1 S_1^{(VM)} + \tau M_2^{V} + \alpha_V V_2^{(M)} - (\lambda_2(t) + \delta_2)S_2^{(VM)} \\
\dot{R}_2 &= \delta_1 R_1 + \gamma I_2 - \delta_2 R_2
\end{aligned}
$$

In older the older age groups ($i = 3, \ldots, 76$), we have the following equations:

$$\begin{aligned}
\dot{V}_i^{(M)} &= \delta_{i-1}V_{i-1}^{(M)} + p_{i-1}(t)\bar{\varepsilon}\delta_{i-1}M_{i-1}^{(V)} + p_{i-1}(t)\bar{\varepsilon}\delta_{i-1}S_{i-1}^{(VM)} - (\alpha_V + \delta_i)V_i^{(M)} \\
\dot{V}_i^{(\bar{M})} &= \delta_{i-1}V_{i-1}^{(\bar{M})} + p_{i-1}(t)\varepsilon\delta_{i-1}S_{i-1}^{(V\bar{M})} - (\alpha_V + \delta_i)V_i^{(\bar{M})} \\
\dot{V}_i &= \delta_{i-1}V_{i-1} + p_{i-1}(t)\varepsilon\delta_{i-1}S_{i-1}^{(V)} - (\alpha_V + \delta_i)V_i \\
\dot{M}_i &= \delta_{i-1}M_{i-1} - (\tau + \delta_i)M_i \\
\dot{M}_i^{(V)} &= \delta_{i-1}M_{i-1}^{(V)} - p_{i-1}(t)\bar{\varepsilon}\delta_{i-1}M_{i-1}^{(V)} - (\tau + \delta_i)M_i^{(V)} \\
\dot{S}_i^{(\bar{M})} &= \delta_{i-1}S_{i-1}^{(\bar{M})} + \tau M_i - (\lambda_i(t) + \delta_i)S_i^{(\bar{M})} \\
\dot{S}_i &= \delta_{i-1}S_{i-1} - (\lambda_i(t) + \delta_i)S_i \\
\dot{E}_i &= \delta_{i-1}E_{i-1} + \lambda_i(t)(S_i + S_i^{(\bar{M})} + S_i^{(V)} + S_i^{(V\bar{M})} + S_i^{(VM)}) - (\sigma + \delta_i)E_i \\
\dot{I}_i &= \delta_{i-1}I_{i-1} + \sigma E_i - (\gamma + \delta_i)I_i \\
\dot{S}_i^{(V)} &= \delta_{i-1}S_{i-1}^{(V)} - p_{i-1}(t)\varepsilon\delta_{i-1}S_{i-1}^{(V)} + \alpha_V V_i - (\lambda_i(t) + \delta_i)S_i^{(V)} \\
\dot{S}_i^{(V\bar{M})} &= \delta_{i-1}S_{i-1}^{(V\bar{M})} - p_{i-1}(t)\varepsilon\delta_{i-1}S_{i-1}^{(V\bar{M})} + \alpha_V V_i^{(\bar{M})} - (\lambda_i(t) + \delta_i)S_i^{(V\bar{M})} \\
\dot{S}_i^{(VM)} &= \delta_{i-1}S_{i-1}^{(VM)} - p_{i-1}(t)\bar{\varepsilon}\delta_{i-1}S_{i-1}^{(VM)} + \alpha_V V_i^{(M)} + \tau M_i^{(V)} - (\lambda_i(t) + \delta_i)S_i^{(VM)} \\
\dot{R}_i &= \delta_{i-1}R_{i-1} + \gamma I_i - \delta_i R_i
\end{aligned}$$

where:

- $1/\sigma$: average latent period
- $1/\gamma$: average infectious period

We implemented a stochastic analogue of this deterministic system using the tau-leap algorithm, with a fixed time step of 1 day.

#### S1.3 Estimation of relative risk of infection following maternal immunization: mathematical equations

To connect the model outputs to empirical estimates of VE [6, 7, 8], we estimated the relative risk (RR) of contracting pertussis in infants from vaccinated mothers relative to that of unvaccinated mothers. The empirical studies from the literature search used two different approaches to obtain this estimate and in our study, we adopted the approach as used in [6, 9], which is the screening method following [10] and which defines the RR as:

$RR = 1 - VE = \frac{PCV}{(1-PCV)} * \frac{(1-PPV)}{PPV}$

In this equation, $PPV$represents the proportion of immunized pregnant mothers in the population and $PCV$ is the proportion of cases in infants born to immunized mothers. RR can take a range of values with (i) 0 indicating no infections in infants of unvaccinated mothers, (ii) a value of 1 reflecting an equal number of infections in infants from vaccinated and unvaccinated mothers, and (iii) any value of above 1 indicating a higher number of infections in infants from vaccinated mothers relative to infants from unvaccinated mothers. Note though that RR is subject to transient dynamics over time and hence the RR value that indicates equal risks of infection between infants from vaccinated versus unvaccinated mothers gets close to 1 only once pertussis transmission in the population is at equilibrium (e.g., Fig. 5, Fig. S2).

In the model, $PPV$, the maternal vaccination coverage is a fixed value, and we show the results at three values, 70% for the baseline value, and 50% and 90% for the sensitivity analyses. These values represent the range observed in empirical studies, with maternal immunization increasing from close to 45% in the first year of implementation to 90% in the last year of monitoring (Table S1 and references therein). Note though that onse study [7] has a long sigmoidal increase in maternal immunization coverage, which for simplicity of the model, we ignore. We expect that, if anything, a long sigmoidal shape of vaccination coverage during the introduction of the maternal immunization program will increase the duration of the transient phase.

$PCV$ represents the ratio of incidence rates, which in newborns is defined as: $\frac{CC^{(\bar{M})}}{CC^{(\bar{M})}+CC^{(S)}}$ , where $CC^{(\bar{M})}$ is the incidence from the $S^{(\bar{M})}$ compartment and $CC^{(S)}$ is the incidence from the $S$ compartment. For any other age class, $PCV$ is defined as: $\frac{CC^{(V\bar{M})}+CC^{VM}}{CC^{(V\bar{M})}+CC^{(VM)}+CC^{(V)}}$, where $CC^{(V\bar{M})}$ is the incidence from the $S^{(V\bar{M})}$ compartment, $CC^{(VM)}$ is the incidence from the $S^{(VM)}$ compartment, and $CC^{(V)}$ is the incidence from the $S^{(V)}$ compartment. We validated the RR estimates of the simulations by setting the duration of maternal derived immunity to infinity, which resulted in 5% RR in newborns or an effectiveness of maternal immunization (VE) of 95% (Fig. S2), which is the initial value set in the simulations (Table S3).

### S2 Supplementary figures

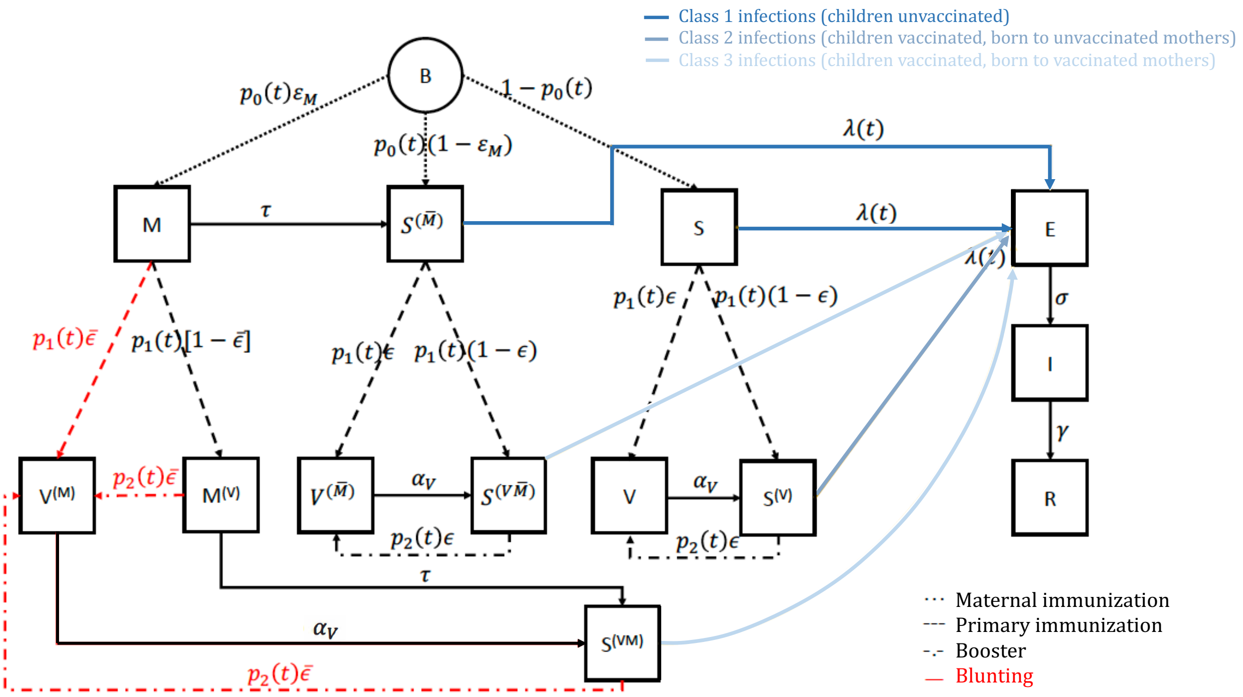


Figure S1: Model schematic with the immunization histories starting from the mothers followed by the infant's primary immunization and booster. Using this schematic, the RR of contracting an infection following maternal immunization can be calculated in the same way as in empirical studies by comparing the incidence of class 2 and class 3 infections. For simplicity, we omitted the transitions between age groups for individuals remaining in the same immunization status, e.g., a fraction $1 - p_1(t)$ transitioning upon aging from $M_1$ to $M_2$ .

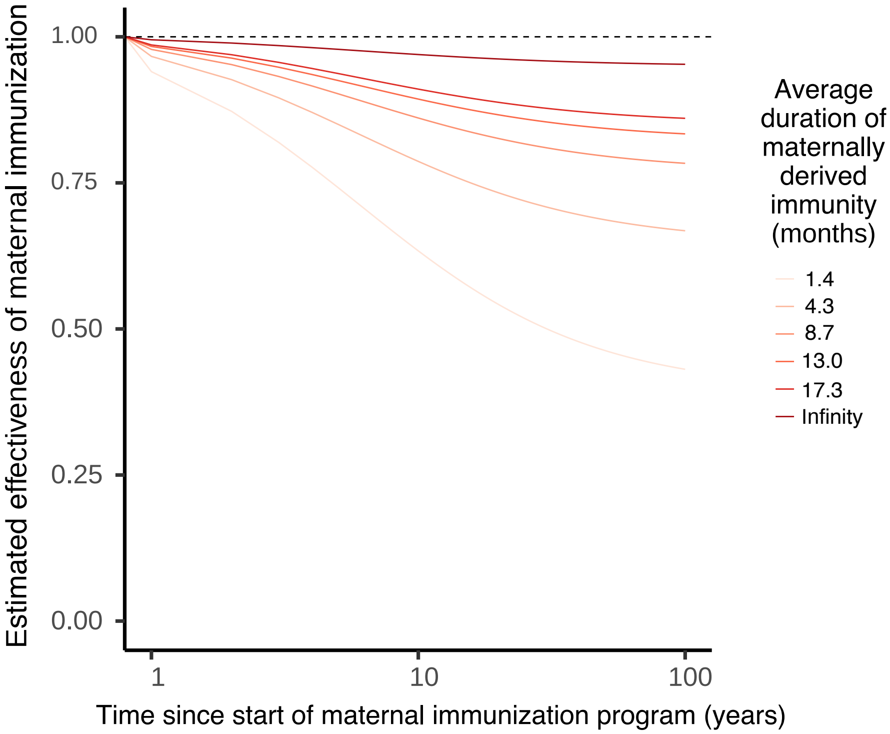


Figure S2: Population-level validation of the estimation of maternal vaccine effectiveness in newborns between the age of 0 and 2 months, with varying duration of maternally derived immunity. In this figure, the dependent variable represents the VE calculated at different time points since the start of maternal immunization (that is, in different cohorts of newborns between 0 and 2 months and is therefore comparable to Fig. 5 in the main text (which represents RR=1–VE in vaccinated infants). In this figure, the gradual decrease in VE over time is caused by the same transient effect as described in the results section: in brief, the deleterious impact of blunting takes years to become manifest, until blunted-vaccinated children reach school age. When the duration of maternally-derived antibodies is assumed infinite, this figure shows that, after the transient phase, the VE estimated from the screening method reaches the nominal value fixed in our model (0.95). When the average duration of maternally derived immunity is close to 8.7 months, which is the value used in the simulations, we obtain at equilibrium, an effectiveness in newborns close to 80%, which is the estimate obtained in some studies, e.g., [11].

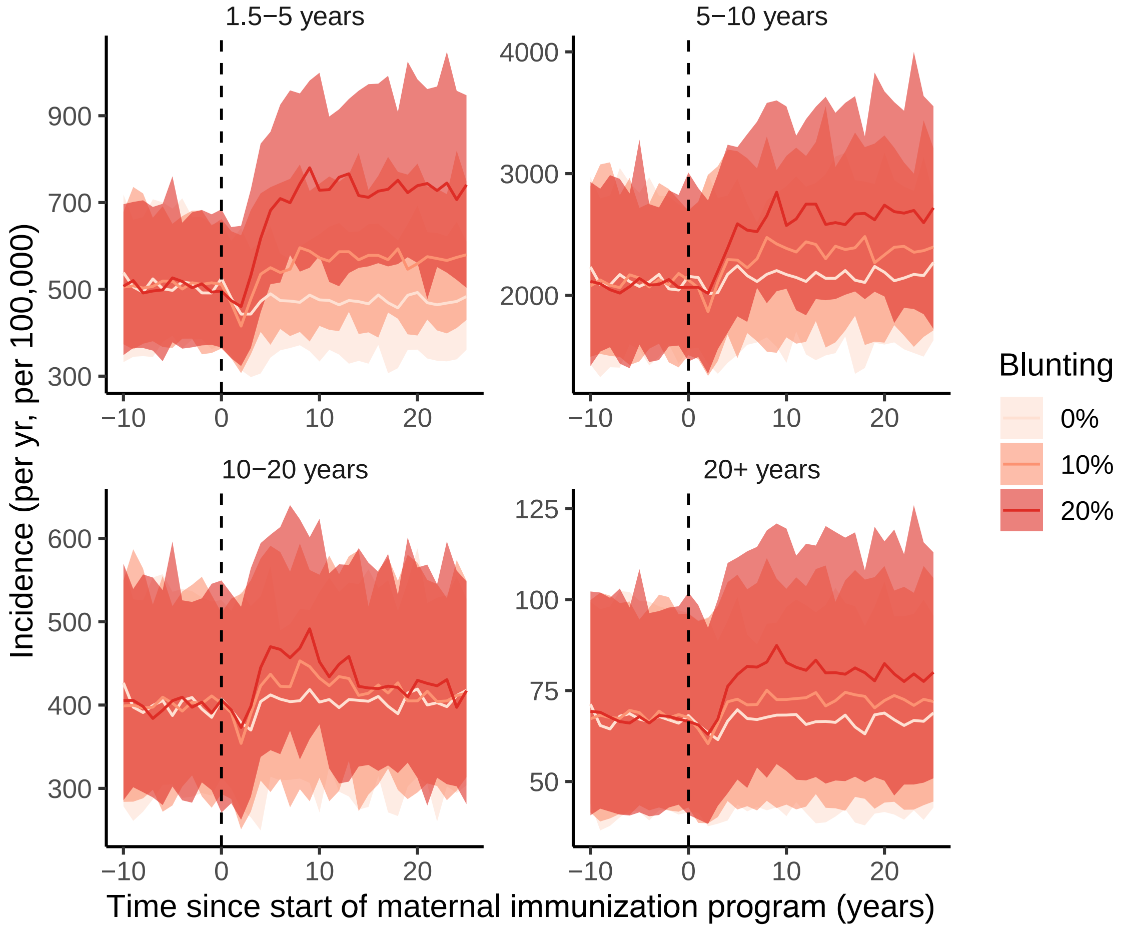


Figure S3: Time series of incidence following maternal immunization with and without blunting across all age classes with maternal immunization coverage at 70%. Blunting refers to a reduction in the effectiveness of the infant primary immunization compared to a non-blunting scenario, for which the effectiveness of the primary immunization is fixed at 96% (Table S3). All else is equal as in Fig. 4.

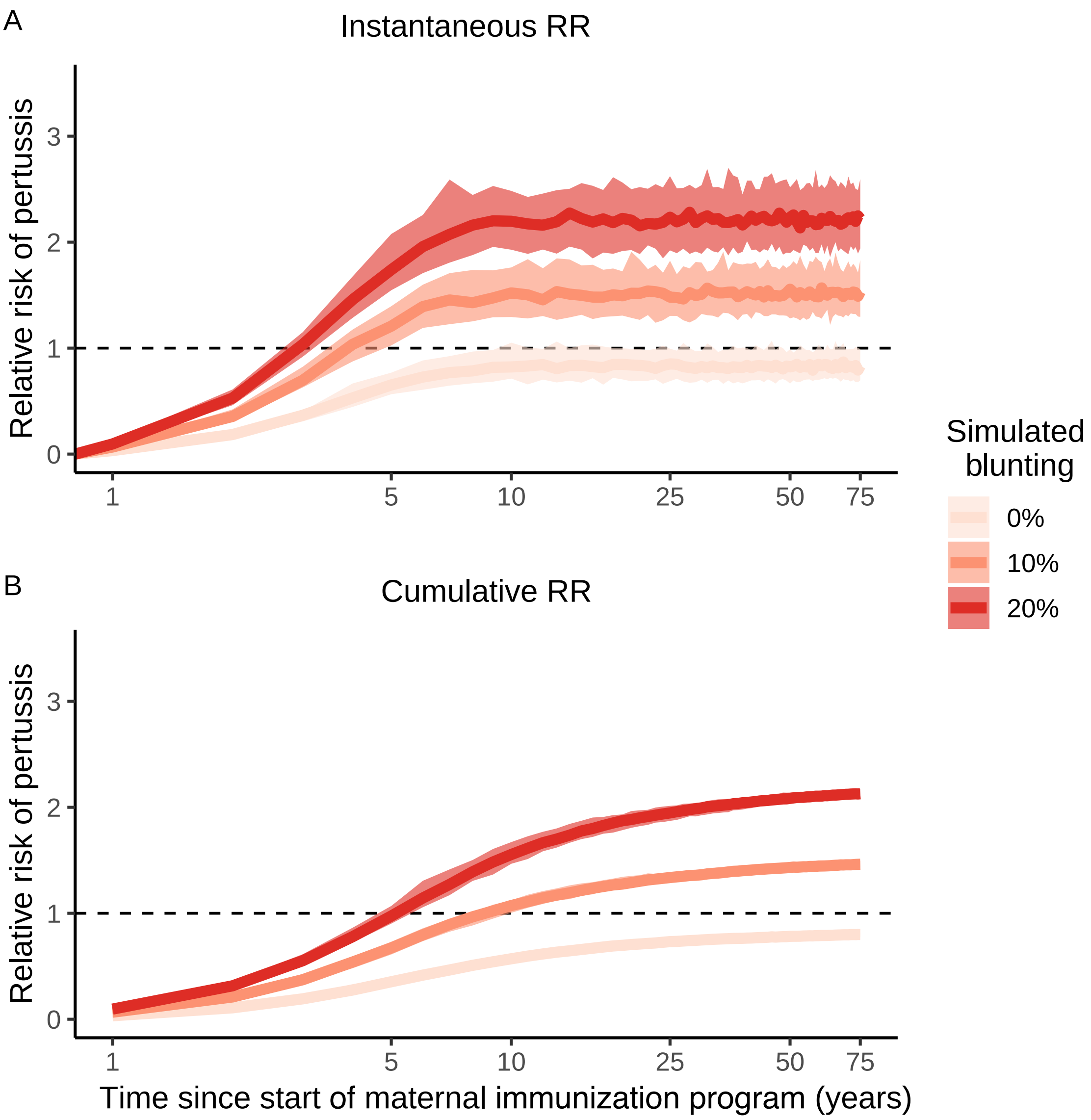


Figure S4: Instant vs. cumulative estimation of RR following maternal immunization after primary immunization, i.e., infants aged 3–18 months. The instantaneous RR was calculated from yearly case counts and the cumulative RR from cumulative yearly case counts since the start of the maternal immunization program. All else is equal as in Fig. 5B.

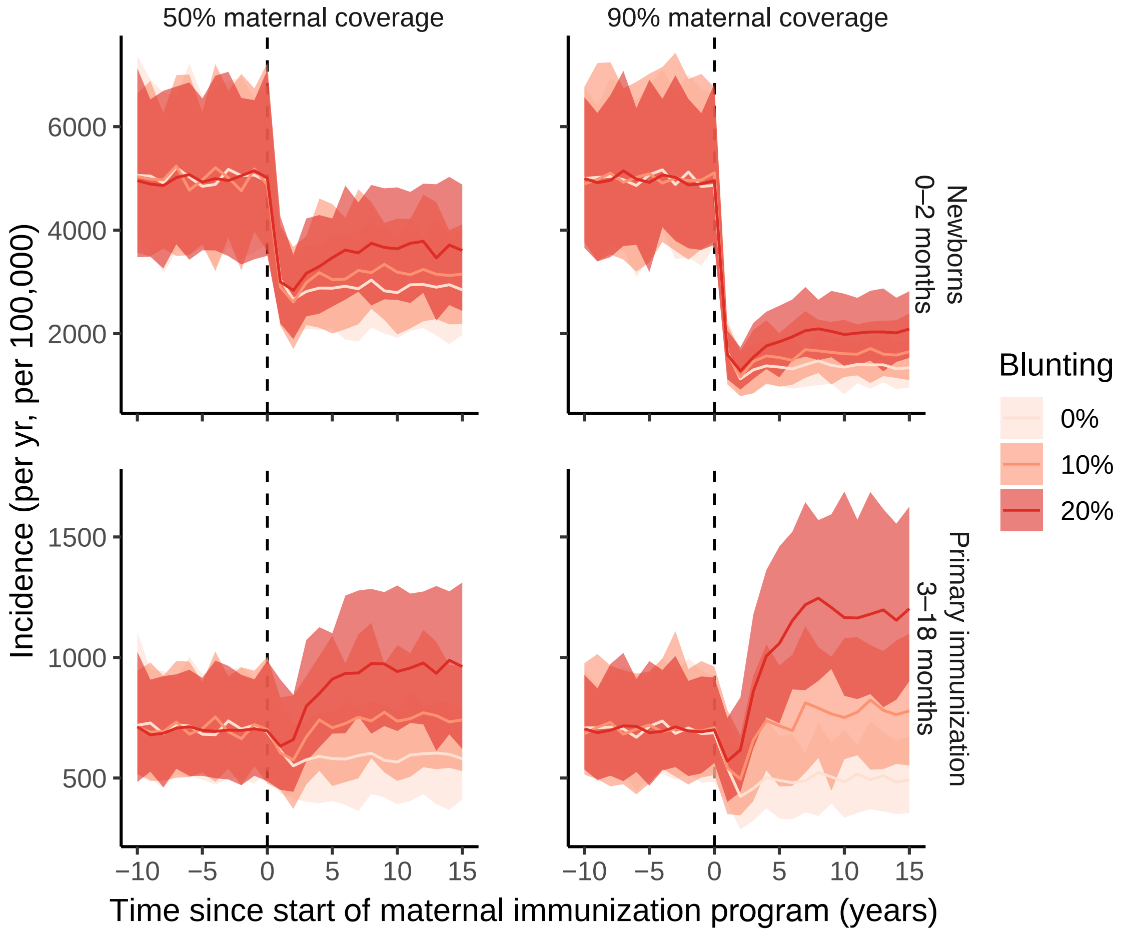


Figure S5: Sensitivity analysis 1: Impact of changing the maternal immunization coverage (base value: 70%, tested values: 50% and 90%). The Y-axis shows the pertussis incidence. Note the different scales of Y-axes between panels. 70% maternal immunization coverage is the value shown in Fig. 4. All else is equal as in Fig. 4.

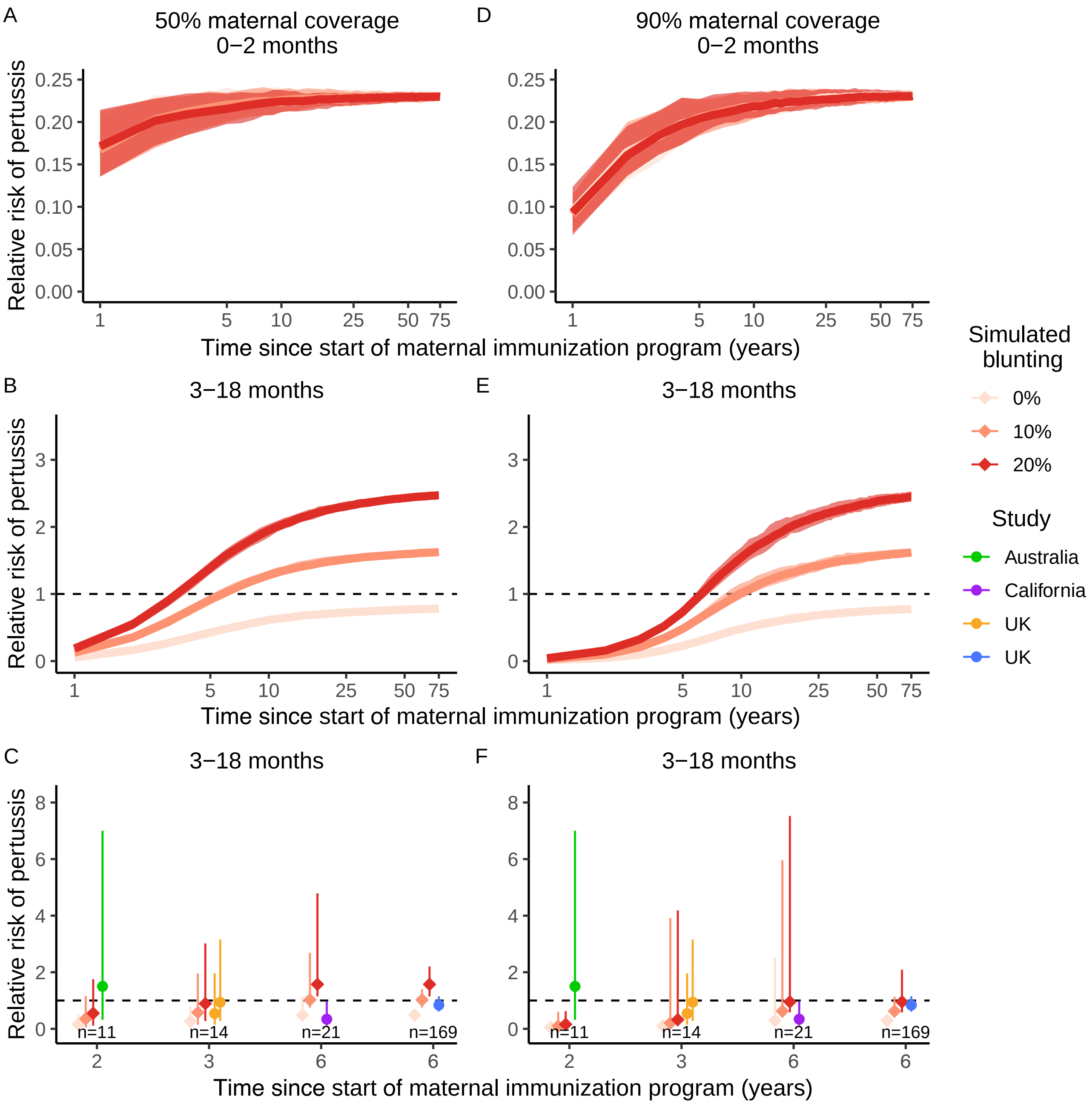


Figure S6: Sensitivity analysis 1: Impact of changing the maternal immunization coverage (base value: 70%, tested values: 50% and 90%). The Y-axis shows the RR of pertussis in infants from vaccinated mothers relative to unvaccinated mothers at (A, B, C) 50% maternal immunization coverage and at (D, E, F) 90% maternal immunization coverage. 70% maternal immunization coverage is the value shown in Fig. 5. All else is equal as in Fig. 5.

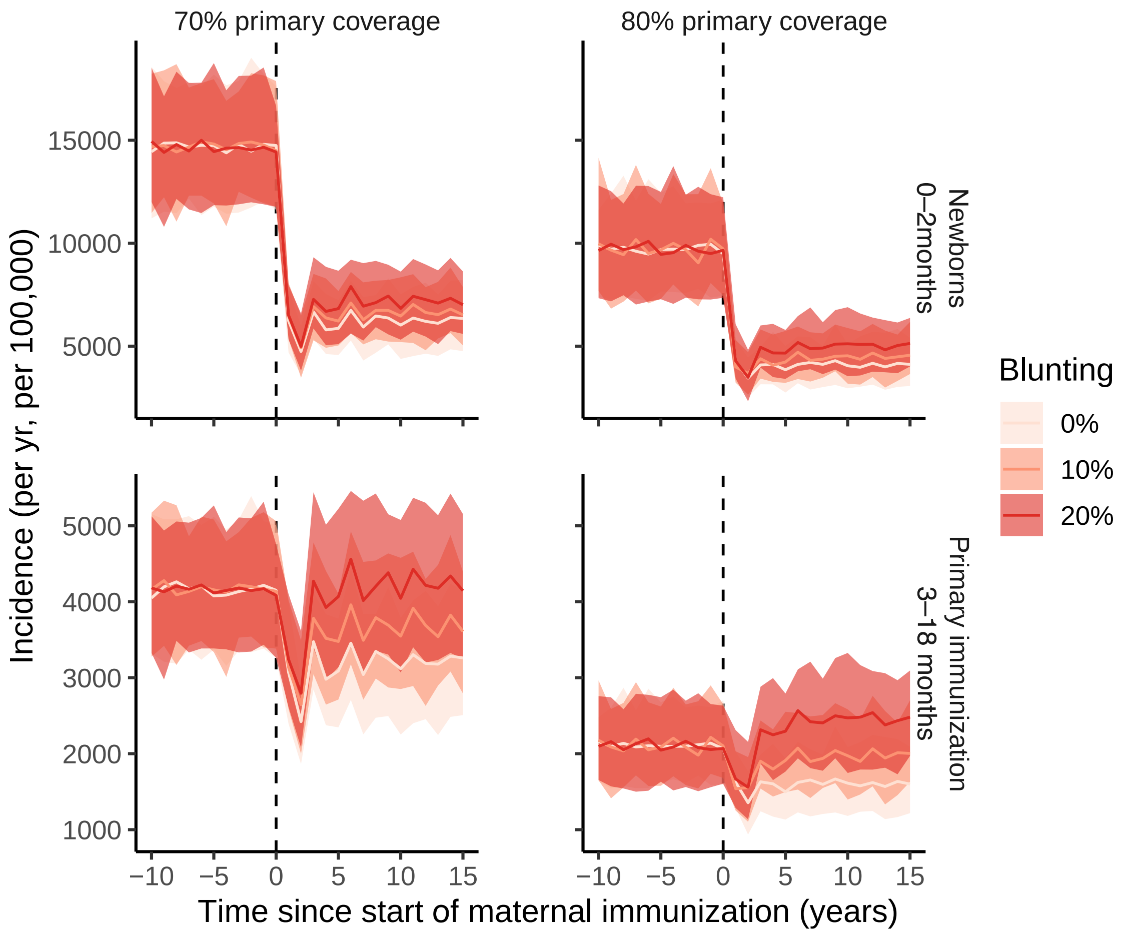


Figure S7: Sensitivity analysis 2: Impact of changing the primary immunization coverage (base value: 90%, tested values: 70% and 80%). The Y-axis shows the pertussis incidence. Note the different scales of the Y-axes between panels. 90% primary immunization coverage is the value shown in Fig. 4. All else is equal as in Fig. 4.

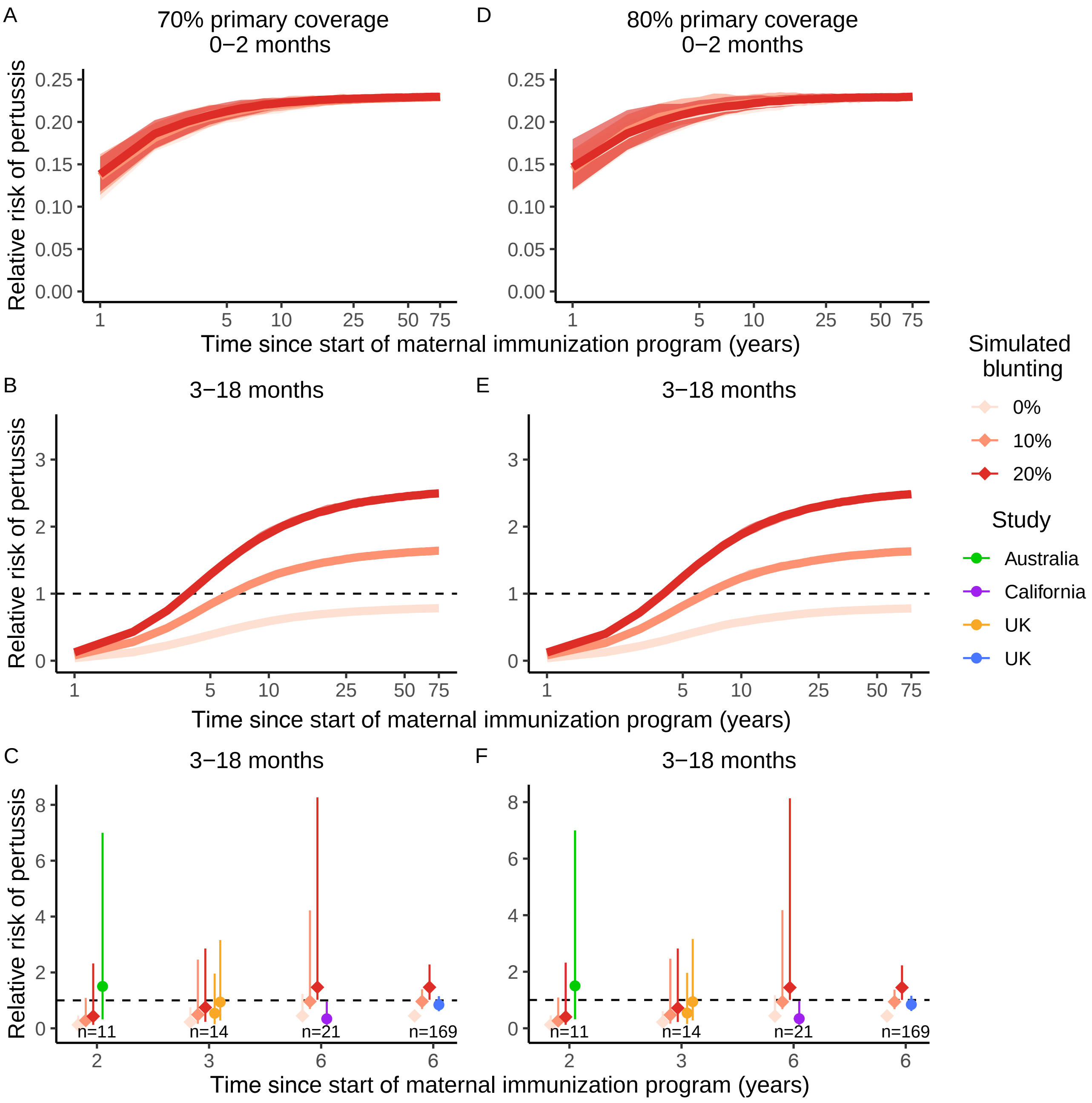


Figure S8: Sensitivity analysis 2: Impact of changing the primary immunization coverage (base value: 90%, tested values: 70% and 80%). The Y-axis shows the RR of pertussis in infants from vaccinated mothers relative to unvaccinated mothers at (A, B, C) 70% primary immunization coverage and at (D, E, F) 80% primary immunization coverage. 90% primary immunization coverage is the value shown in Fig. 5. All else is equal as in Fig. 5.

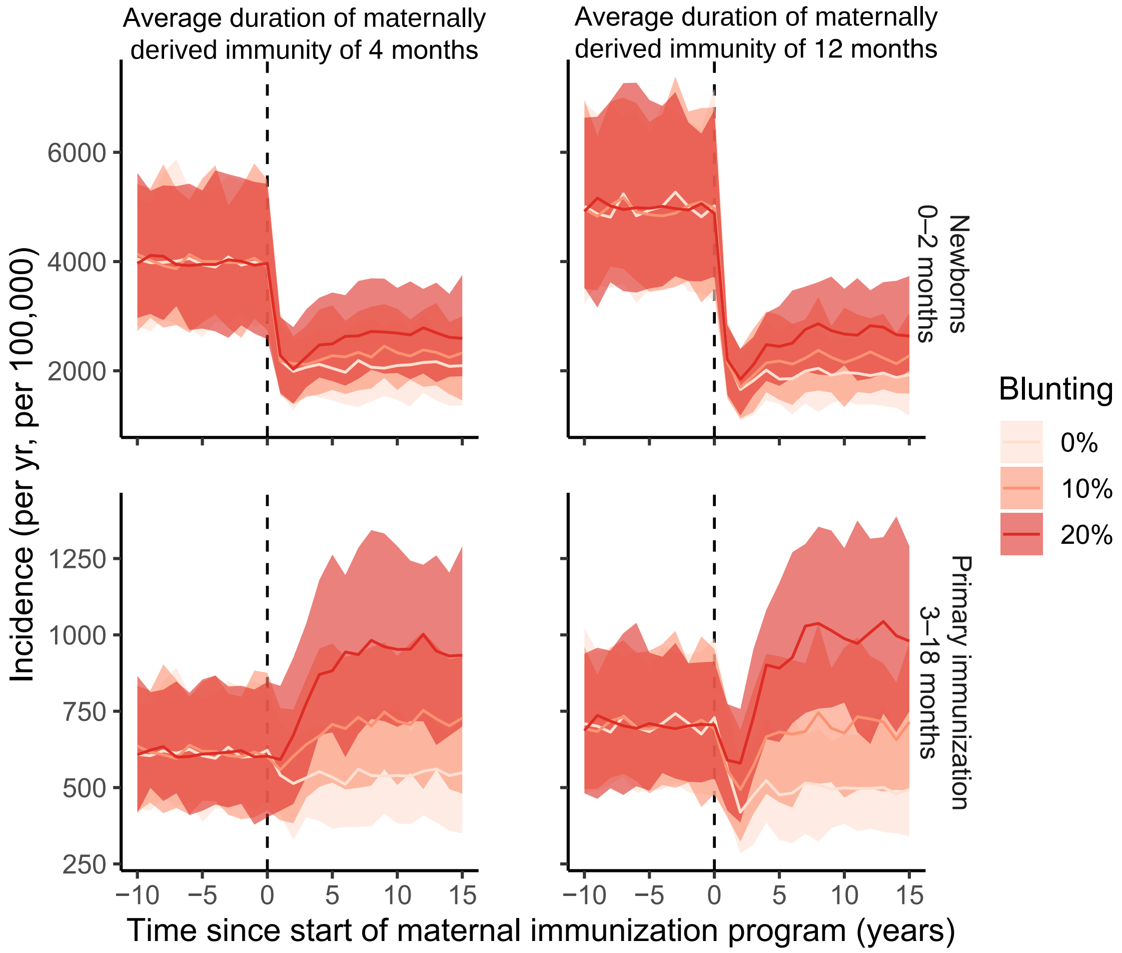


Figure S9: Sensitivity analysis 3: Impact of changing the average duration of maternally derived protection (base value: 8.7 months, tested values: 4 months and 12 months). The Y-axis shows the pertussis incidence. Note the different scales of the Y-axes between panels. 8.7 months is the value shown in Fig. 4. All else is equal as in Fig. 4.

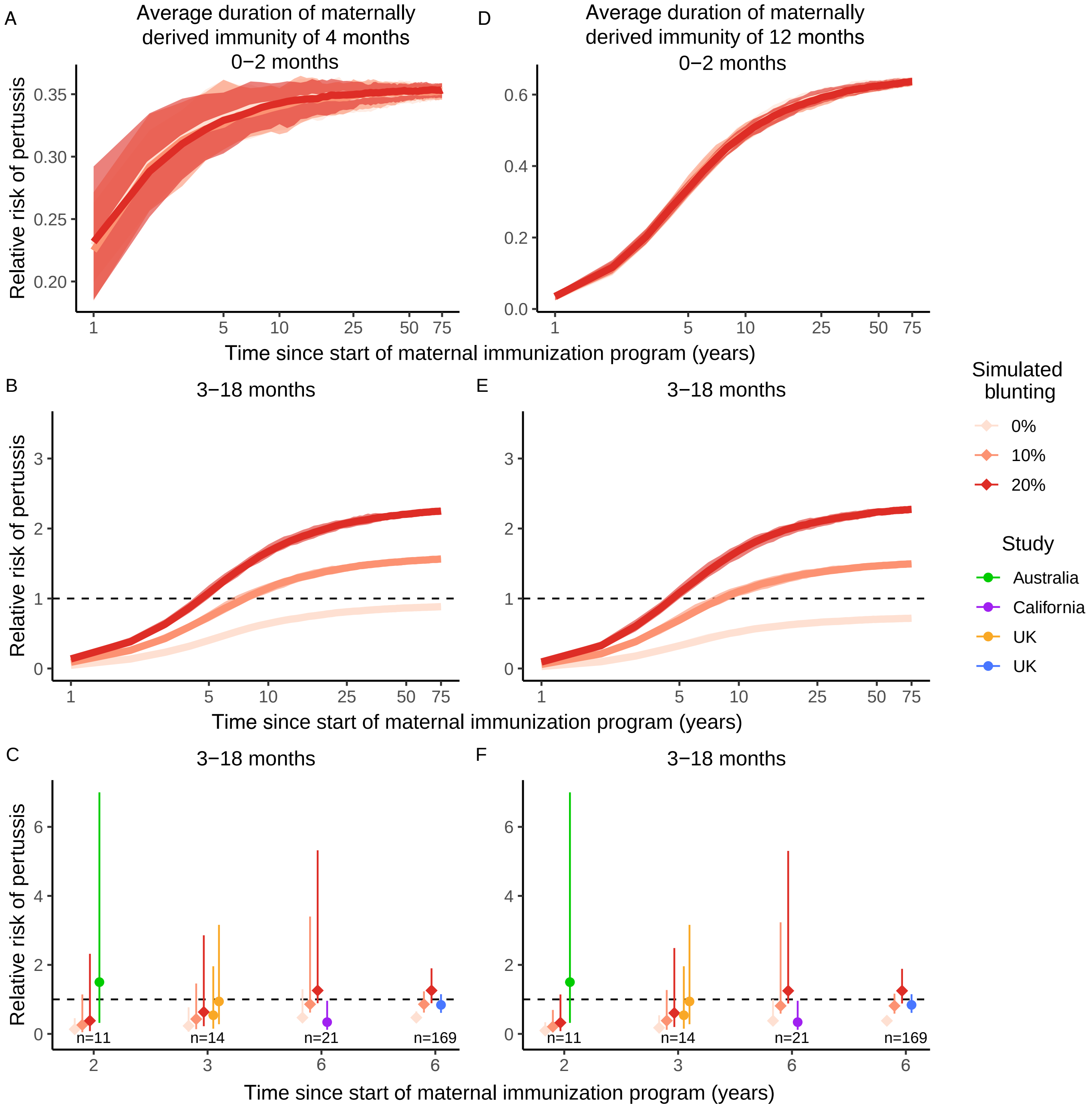


Figure S10: Sensitivity analysis 3: Impact of changing the average duration of maternal protection (base value: 8.7 months, tested values: 4 months and 12 months). The Y-axis shows the RR of pertussis in infants from vaccinated mothers relative to unvaccinated mothers when the average duration of maternally derived protection is either (A, B, C) 4 months or (D, E, F) 12 months. Note the different scales on the Y-axes between panels. 8.7 months is the value shown in Fig. 5. All else is equal as in Fig. 5.

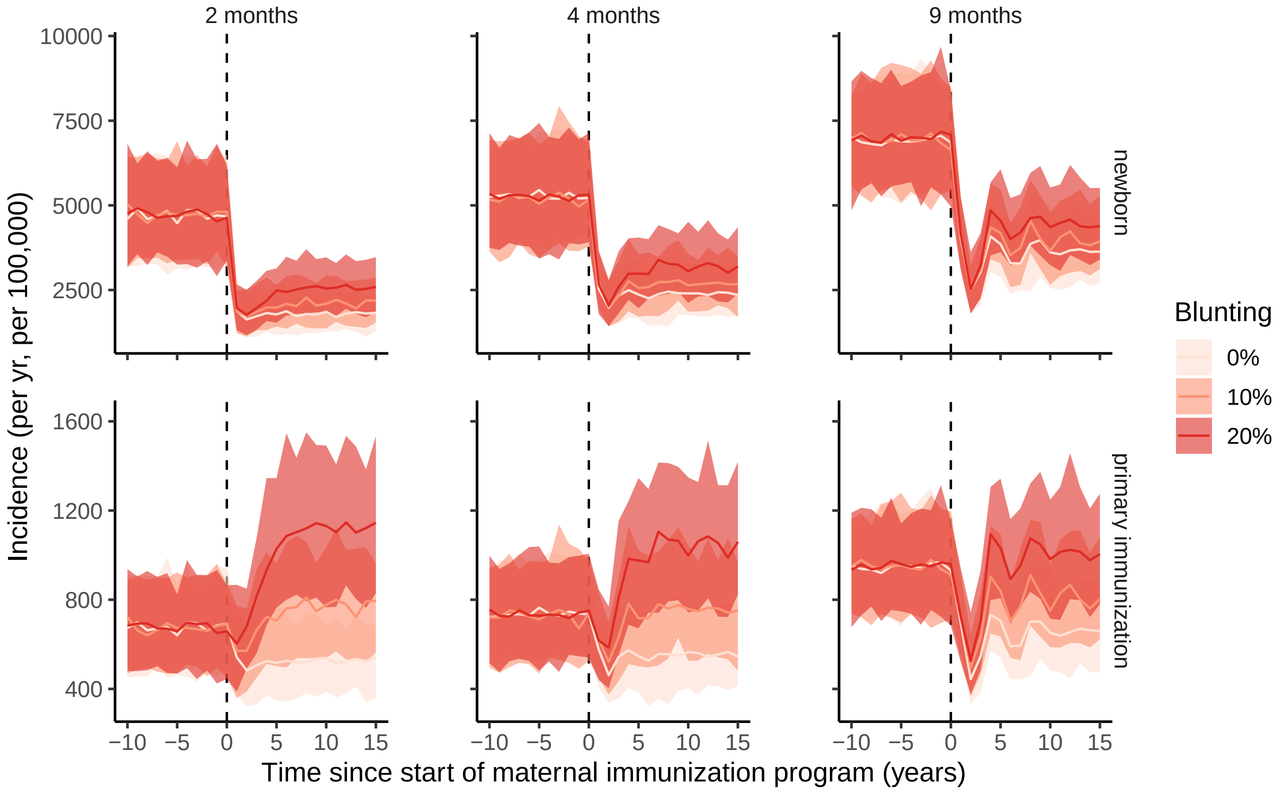


Figure S11: Sensitivity analysis 4: Impact of changing the age at start of the infant primary immunization (base value: 3 months, tested values: 2, 4 and 9 months). The Y-axis shows the pertussis incidence. Note the different scales of Y-axes between the top and the lower panels. Three months is the value used for Fig. 4. All else is equal as in Fig. 4.

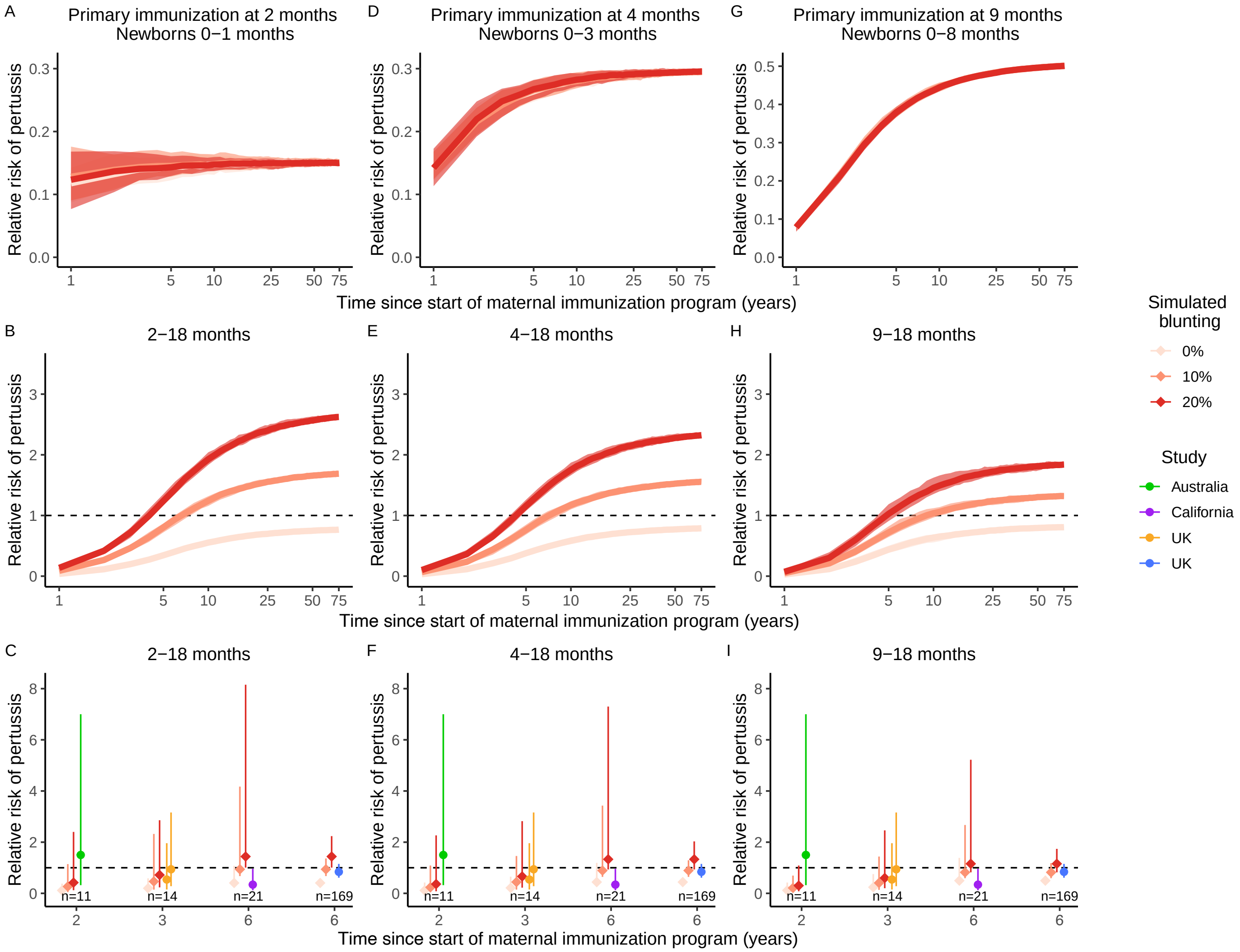


Figure S12: Sensitivity analysis 4: Impact of changing the age at start of the infant primary immunization (base value: 3 months, tested values: 2, 4 and 9 months). The Y-axis shows the RR of pertussis in infants from vaccinated mothers relative to unvaccinated mothers. Three months is the value used for Fig. 5. All else is equal as in Fig. 5.

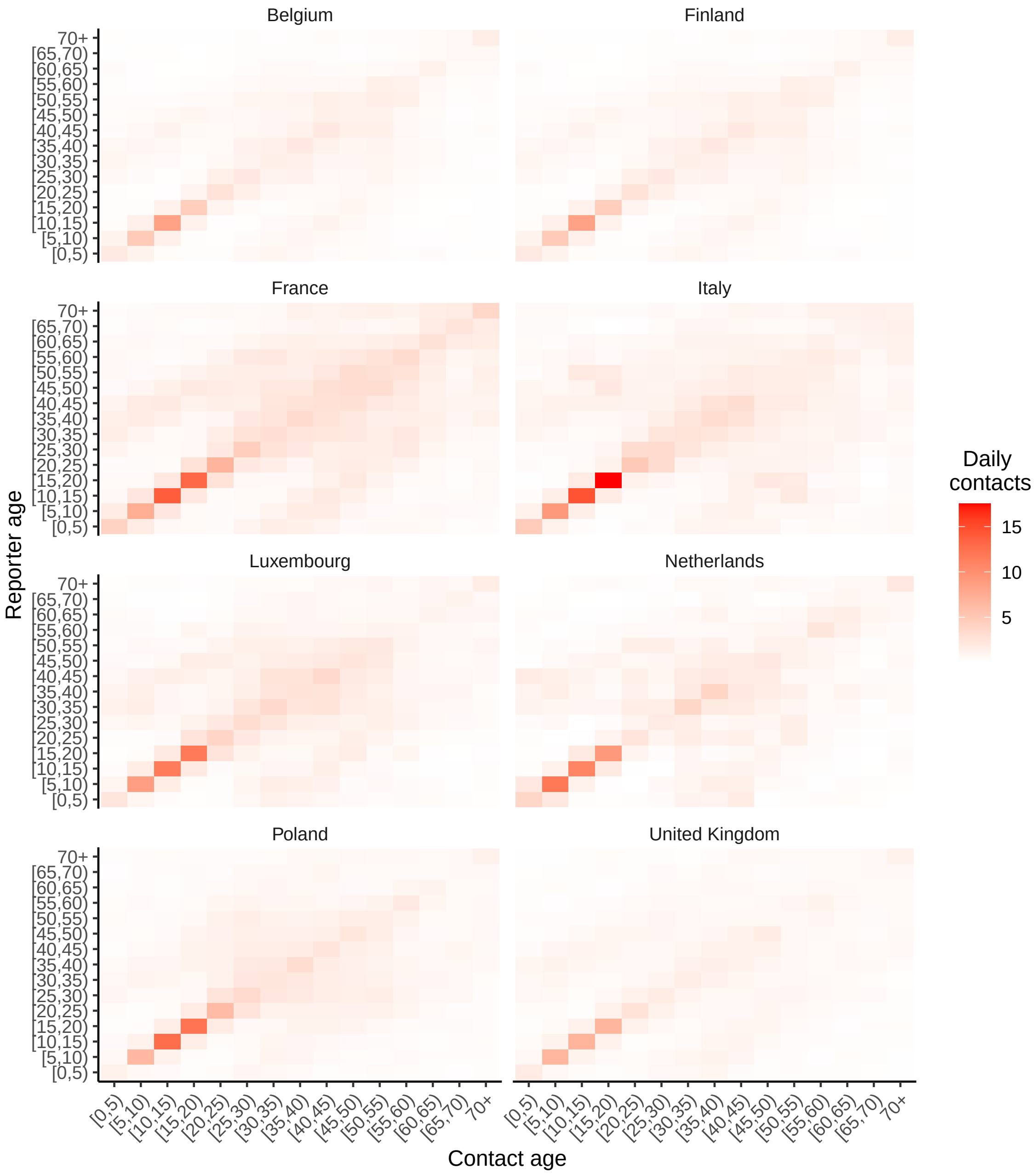


Figure S13: Sensitivity analysis 5: Contact matrices used in the sensitivity analyses for Fig. S14 to Fig. S18. Data for matrices were obtained from [12] and [13] using the package socialmixR [14]. All matrices were corrected for reciprocity. Note that (i) some countries have a larger number of daily contacts between the age groups [5,10), [10,15) and [15,20) relative to other age groups, and (ii) some countries have a larger mixing of [0,5) with adolescents or adults (i.e., light red squares parallel to the same-age diagonal squares). Separate contact information for the under 1 year old was not available from the surveys in [12] and [13].

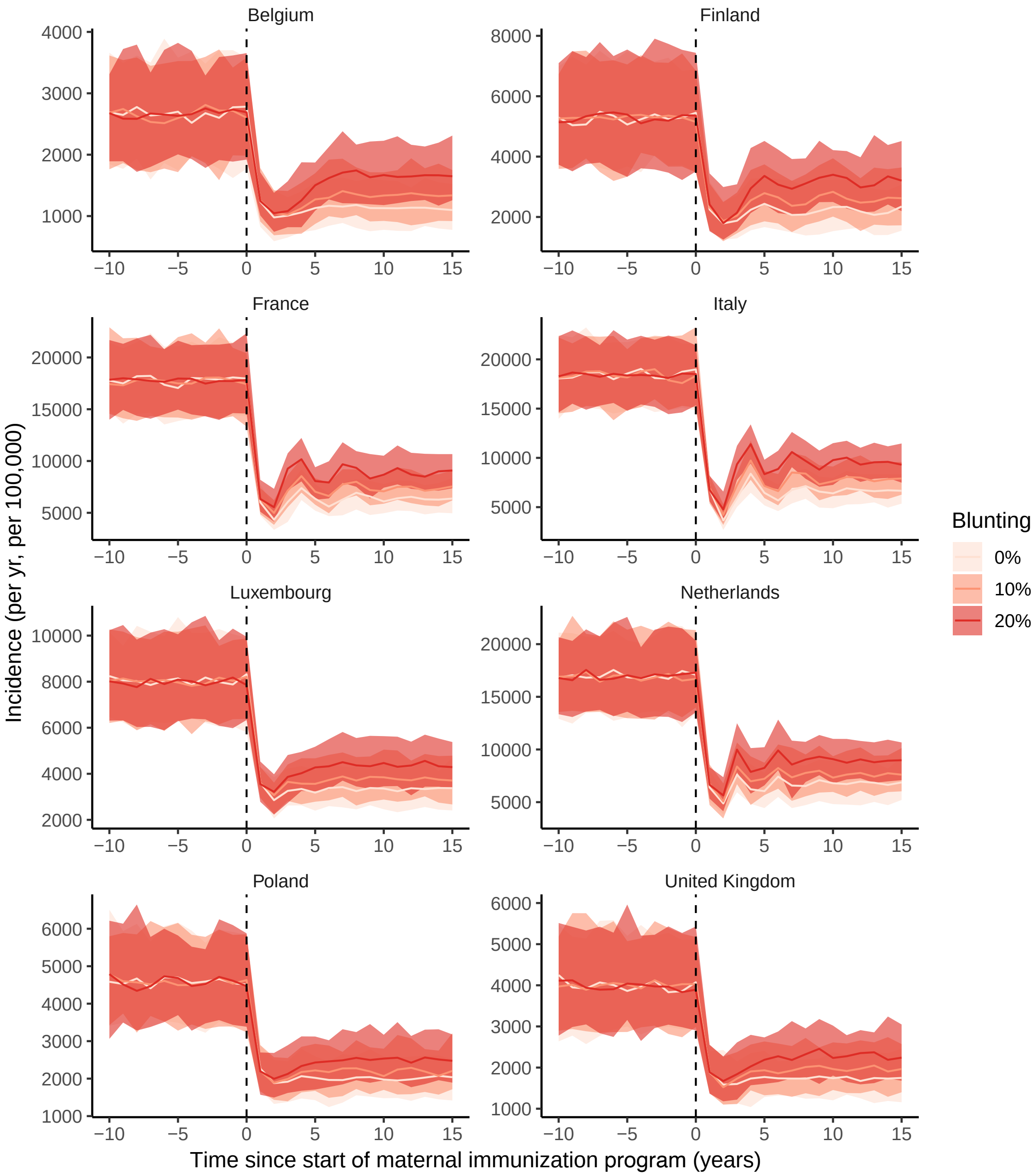


Figure S14: Sensitivity analysis 5: The impact of the contact matrix on pertussis incidence in newborns (0 – 2 months). All panels show a transient phase with low incidence and a rebound. The United Kingdom is the contact matrix used in Fig. 4. Note the different scales on the Y-axes between panels. All else is equal as in Fig. 4.

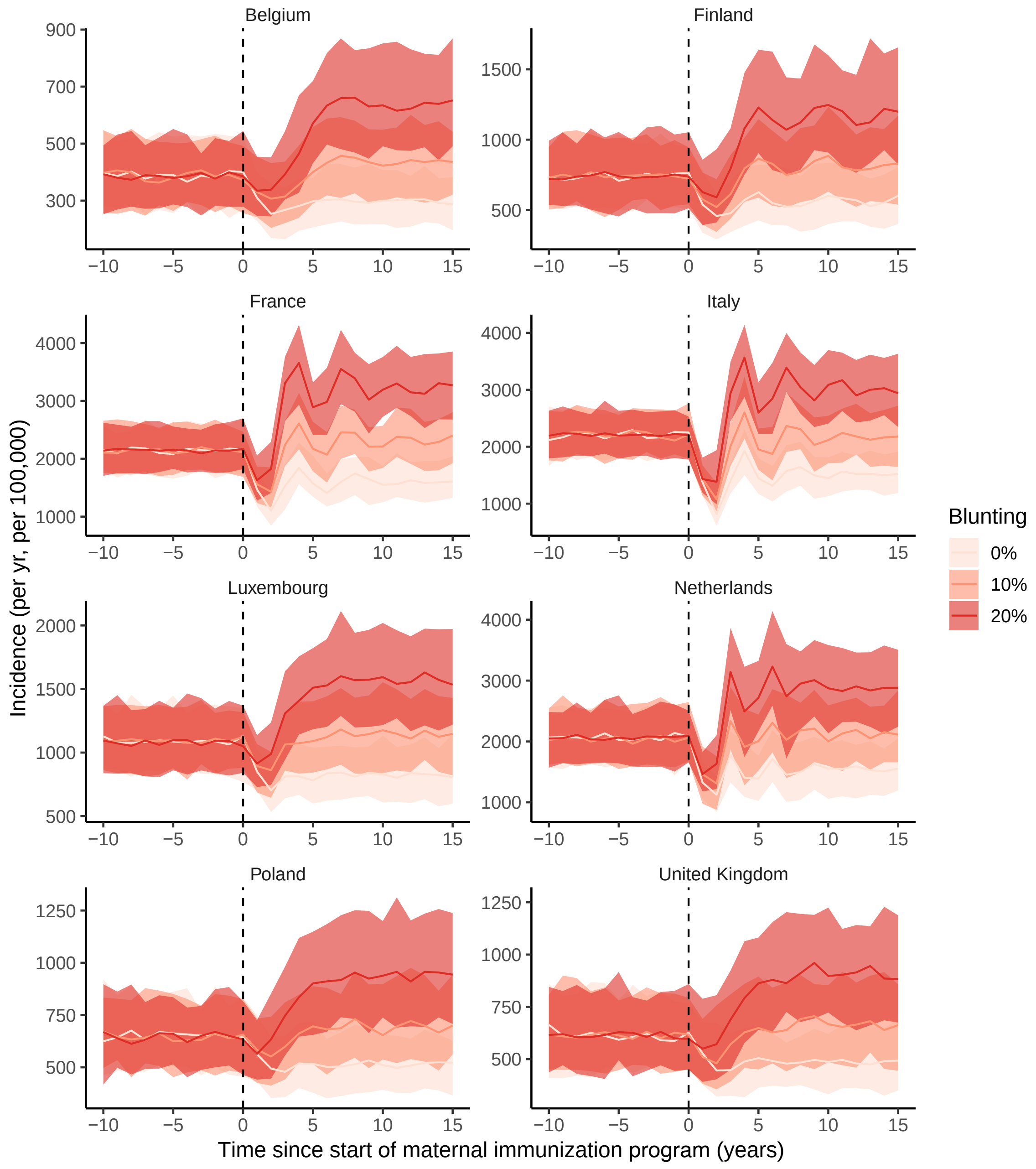


Figure S15: Sensitivity analysis 5: the impact of the contact matrix on pertussis incidence after the primary immunization (between 3 – 18 months). All panels show a transient phase with low incidence and a rebound. The United Kingdom is the contact matrix used in Fig. 4. Note the different scales on the Y-axes between panels. All else is equal as in Fig. 4.

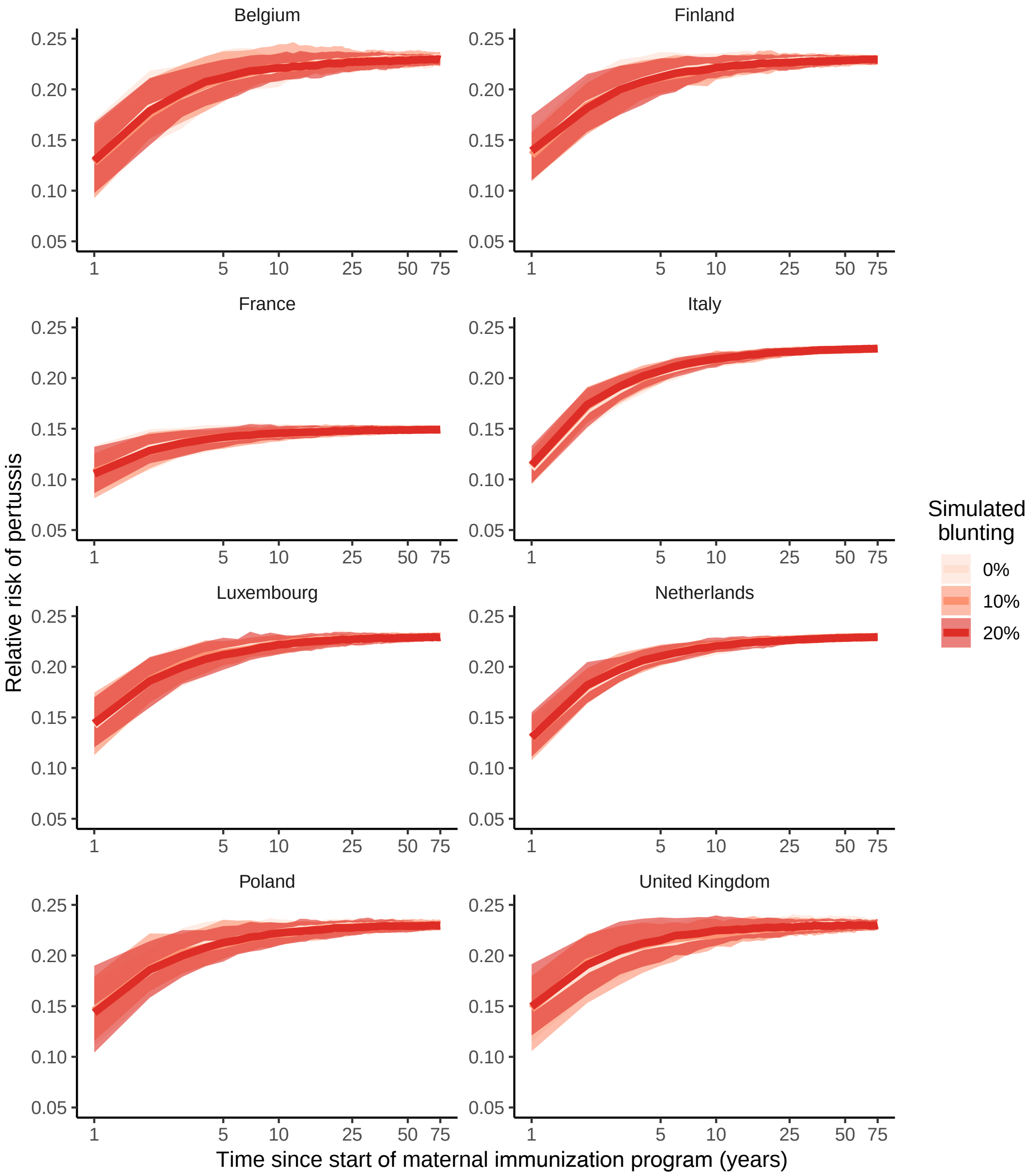


Figure S16: Sensitivity analysis 5: The impact of the contact matrix on the risk of pertussis in newborns (0 – 2 months) from vaccinated mothers relative to unvaccinated mothers. All panels show a transient phase in the RR, with some variation in the duration of this transient phase between countries. The United Kingdom is the contact matrix used in Fig. 5A. All else is equal as in Fig. 5A.

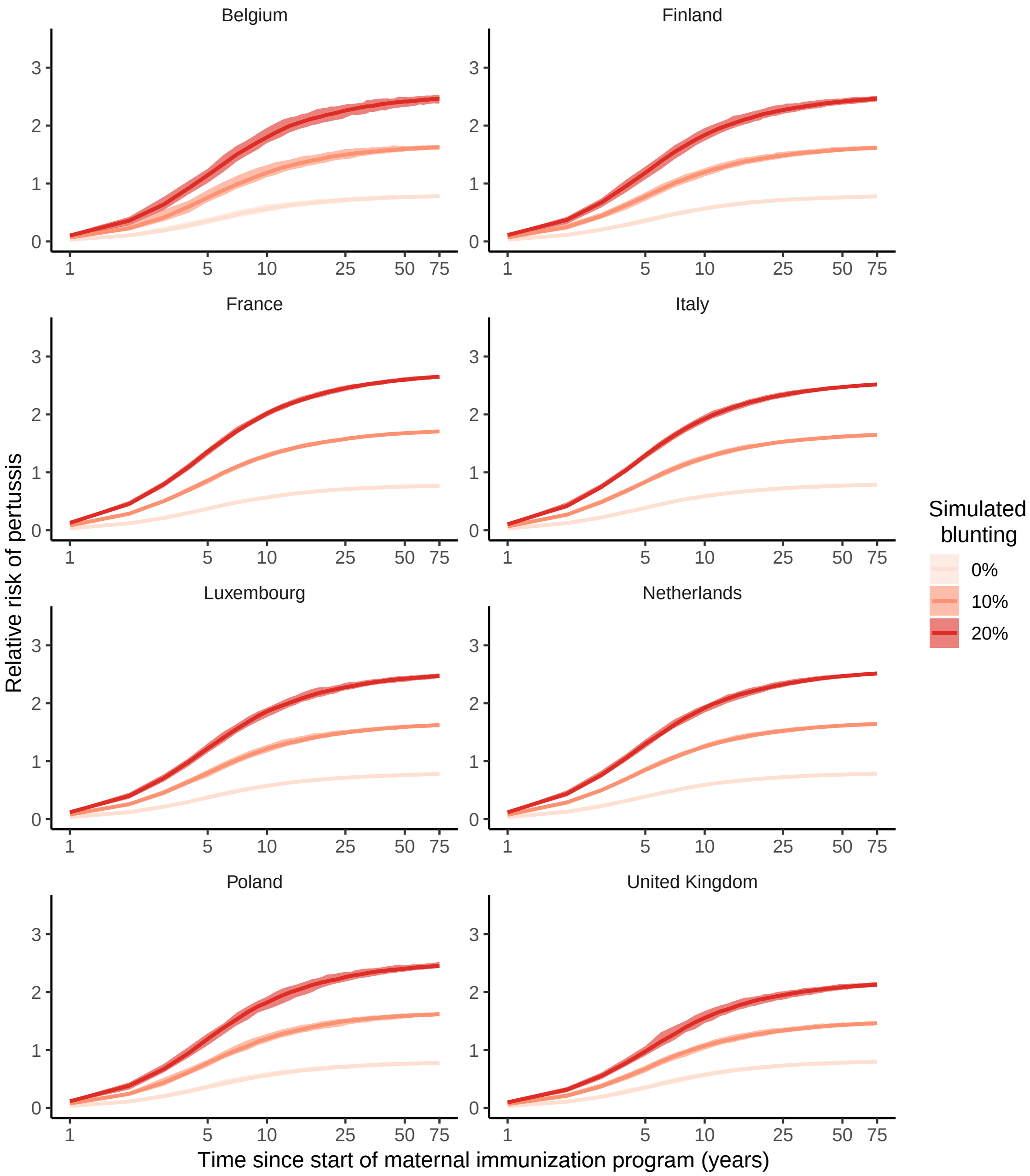


Figure S17: Sensitivity analysis 5: The impact of different contact matrices on the risk of pertussis after the primary immunization (3 – 18 months) in infants from vaccinated mothers relative to unvaccinated mothers. The United Kingdom is the contact matrix used in Fig. 5B. All else is equal as in Fig. 5B.

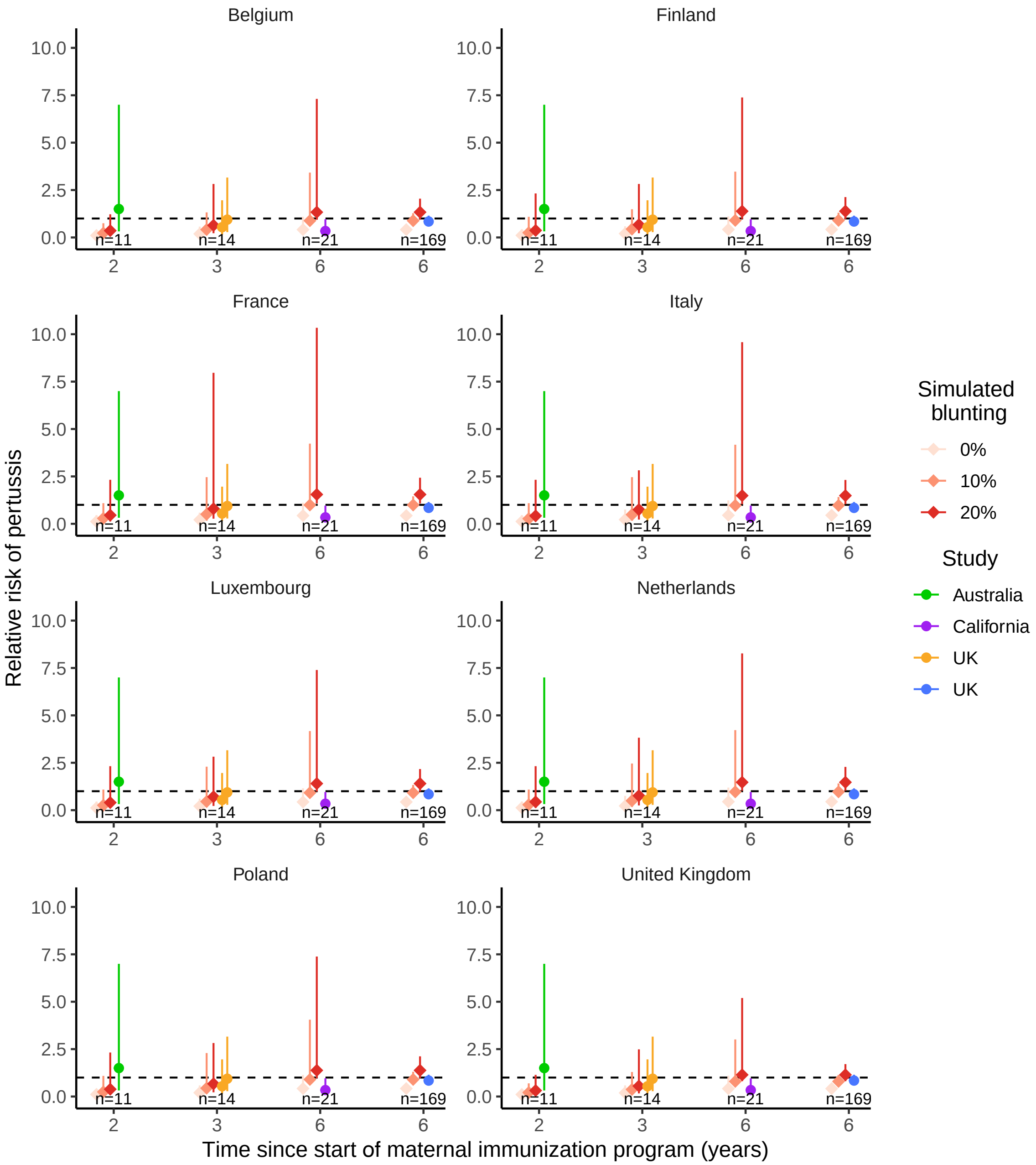


Figure S18: Sensitivity analysis 5: The impact of different contact matrices on the risk of pertussis after the primary immunization (3 – 18 months) in infants from vaccinated mothers relative to unvaccinated mothers. In the simulated estimates, error bars represent 95% CI and are the result of uncertainty due to simulation stochasticity and due to sampling, with sample sizes consistent with those from empirical studies (Table S1). The United Kingdom is the contact matrix used in Fig. 5C. All else is equal as in Fig. 5C.

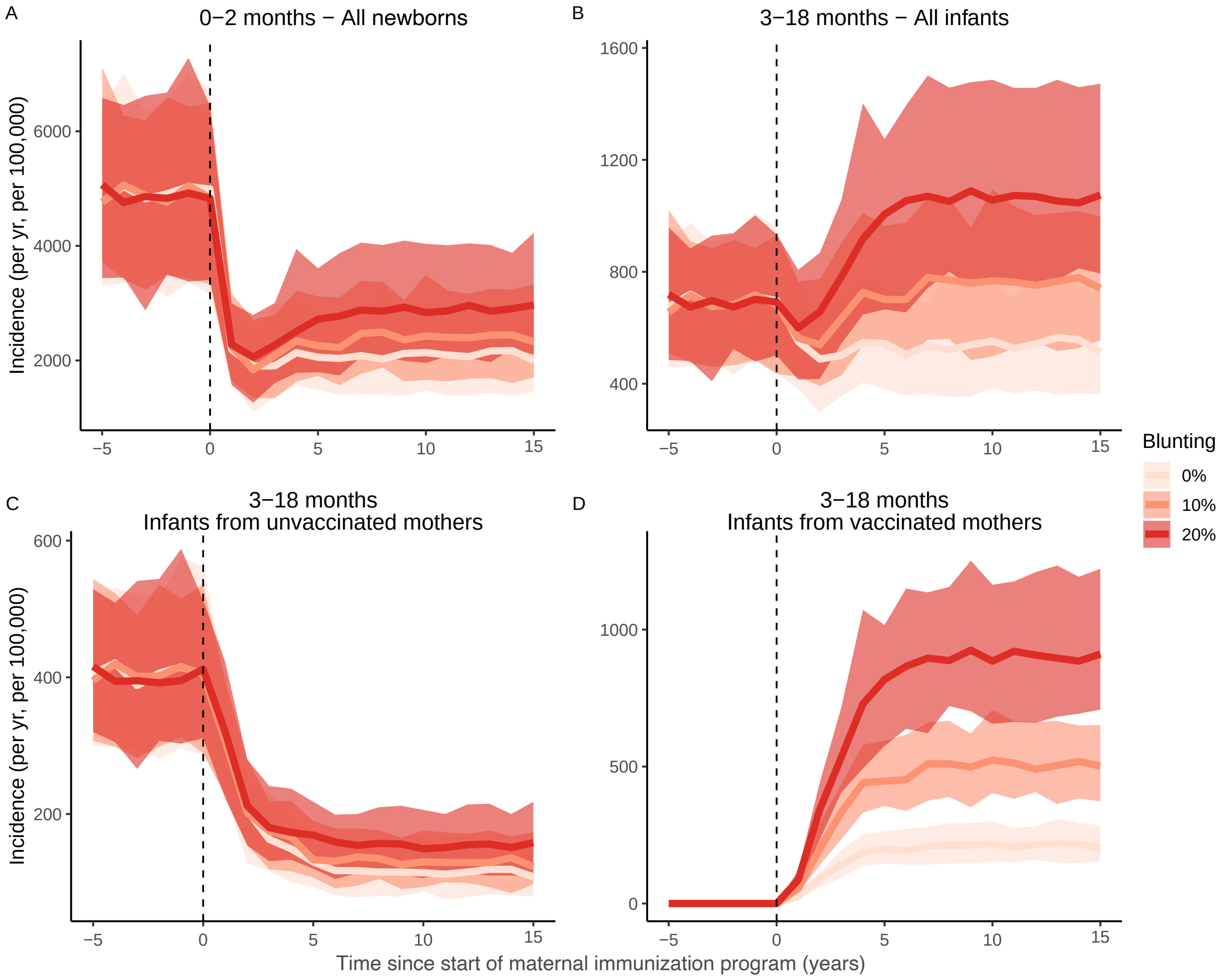


Figure S19: Sensitivity analysis 6: Pertussis incidence with the introduction of the maternal immunization program 60 years after the introduction of the infant immunization program. All other parameter values are as in Fig. 4, for which the maternal immunization program started 100 years after the introduction of the infant immunization program. All else is equal as in Fig. 4.

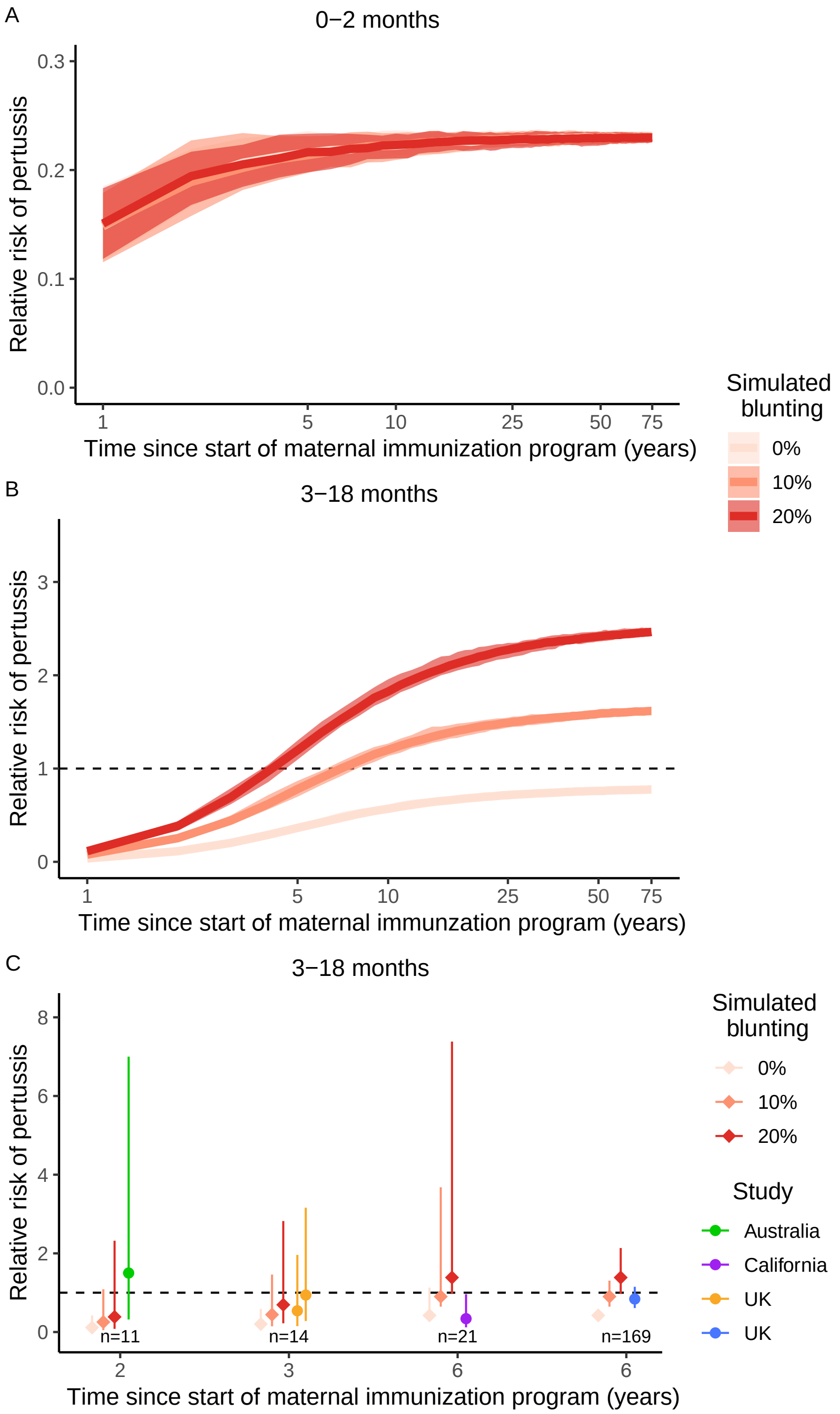


Figure S20: Sensitivity analysis 6: The RR with the introduction of the maternal immunization program 60 years after the introduction of the infant immunization program. All other parameter estimates are as in Fig. 5, for which the maternal immunization program started 100 years after the introduction of the infant immunization program. All else is equal as in Fig. 5.

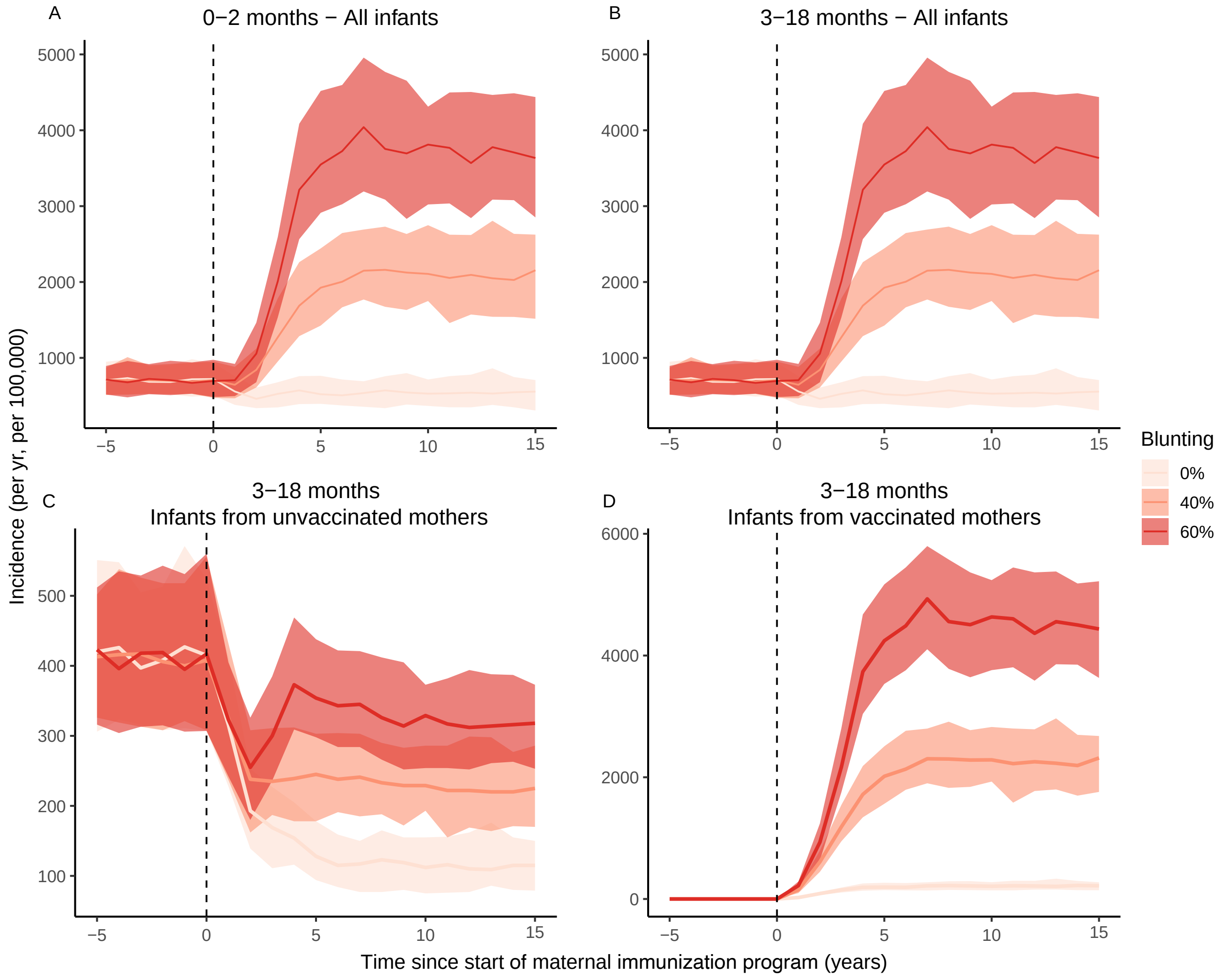


Figure S21: Sensitivity analysis 7: Pertussis incidence before and after the start of the maternal immunization program without blunting (0%) and with blunting levels up to 60%. All other parameter values are as in Fig. 4, which shows scenarios with a maximum of 20% blunting. All else is equal as in Fig. 4.

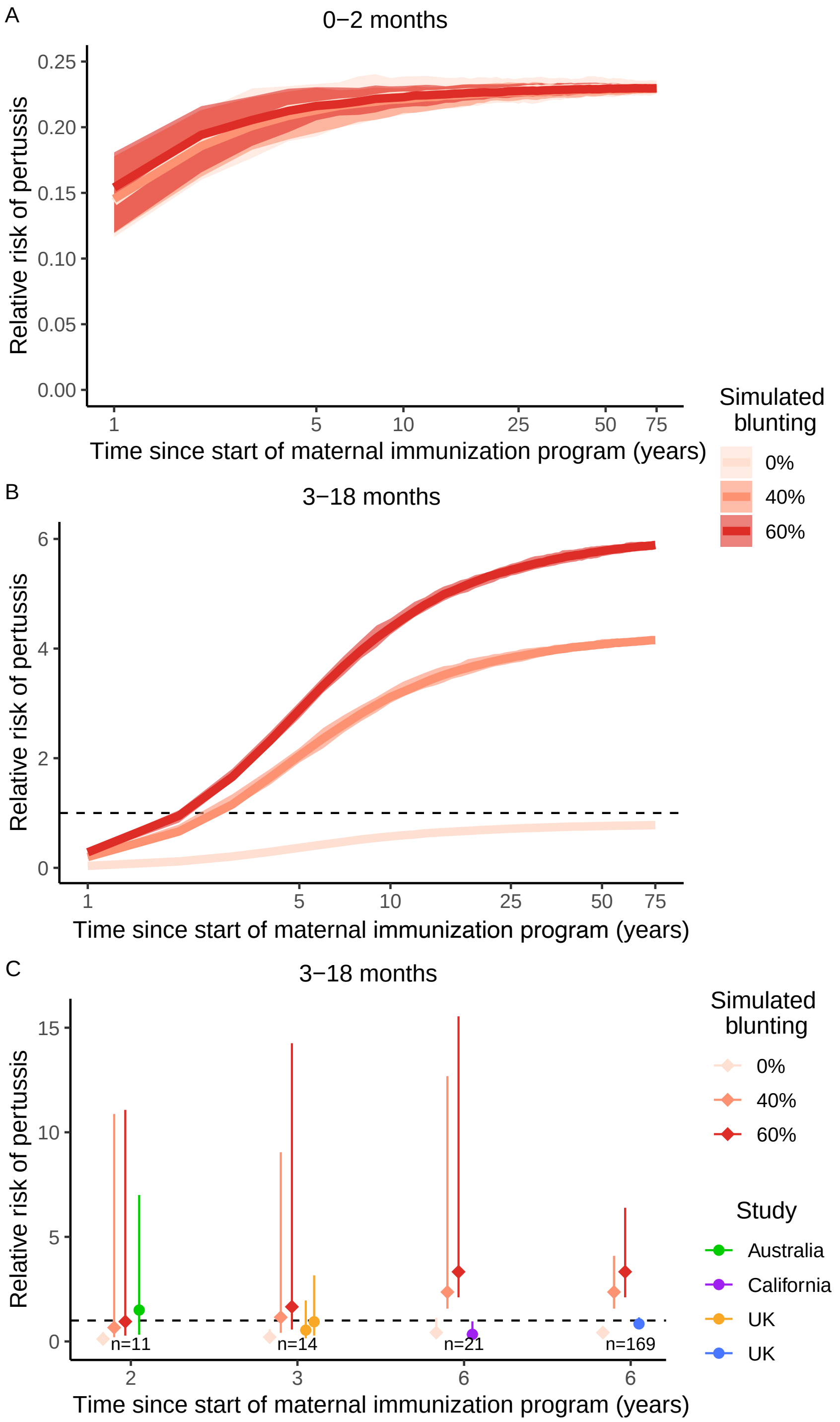


Figure S22: Sensitivity analysis 7: The RR of pertussis infection in infants from vaccinated relative to unvaccinated mothers without blunting (0% blunting) and with blunting levels up to 60% All other parameter values are as in Fig. 5, which shows scenarios with a maximum of 20% blunting. All else is equal as in Fig. 5.

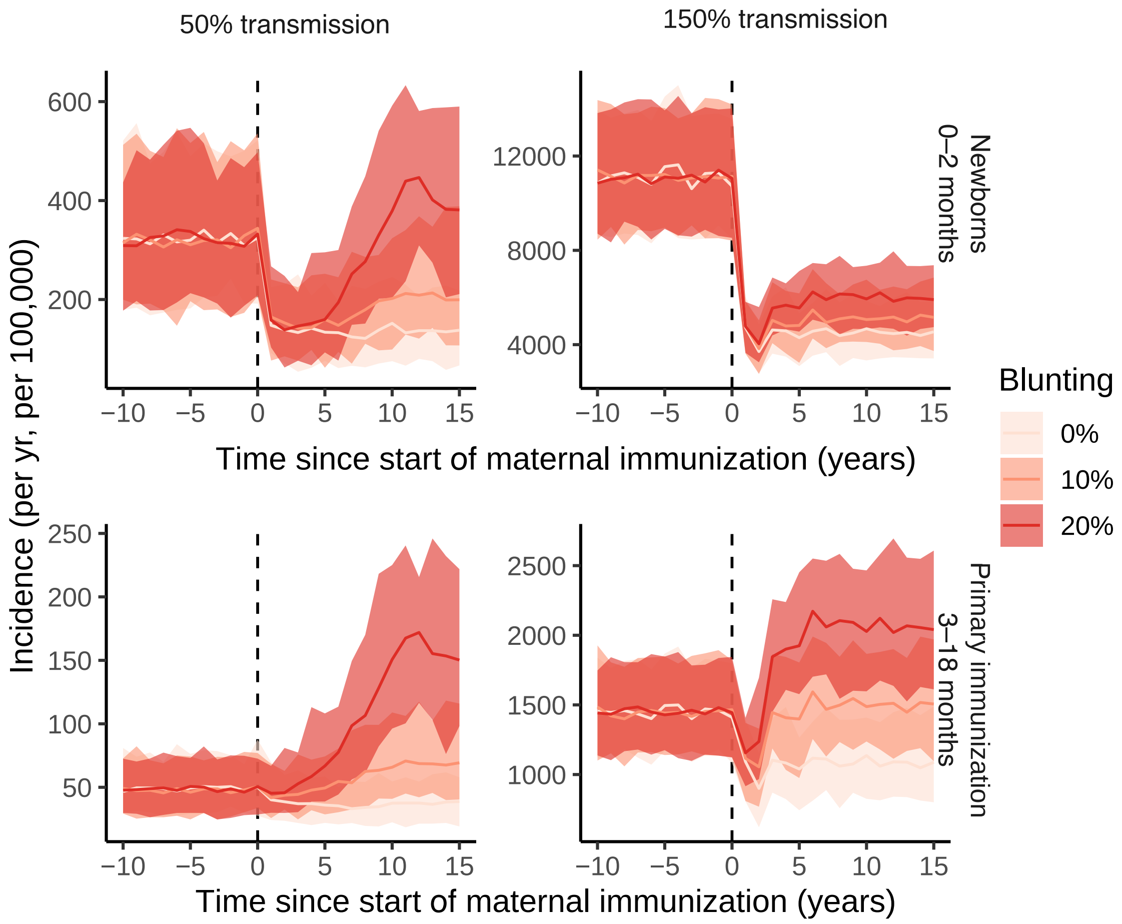


Figure S23: Sensitivity analysis 8: Pertussis incidence before and after the start of the maternal immunization program in a population with the transmission in the 0 to 9-year-old being either low ($q_{1,\ldots,11}$=0.045) or high ($q_{1,\ldots,11}$=0.135). The baseline value in Fig. 4 is 0.09, which was the best-fitting value of the stochastic model on Massachusetts data [1]. Everything else is equal as in Fig. 4.

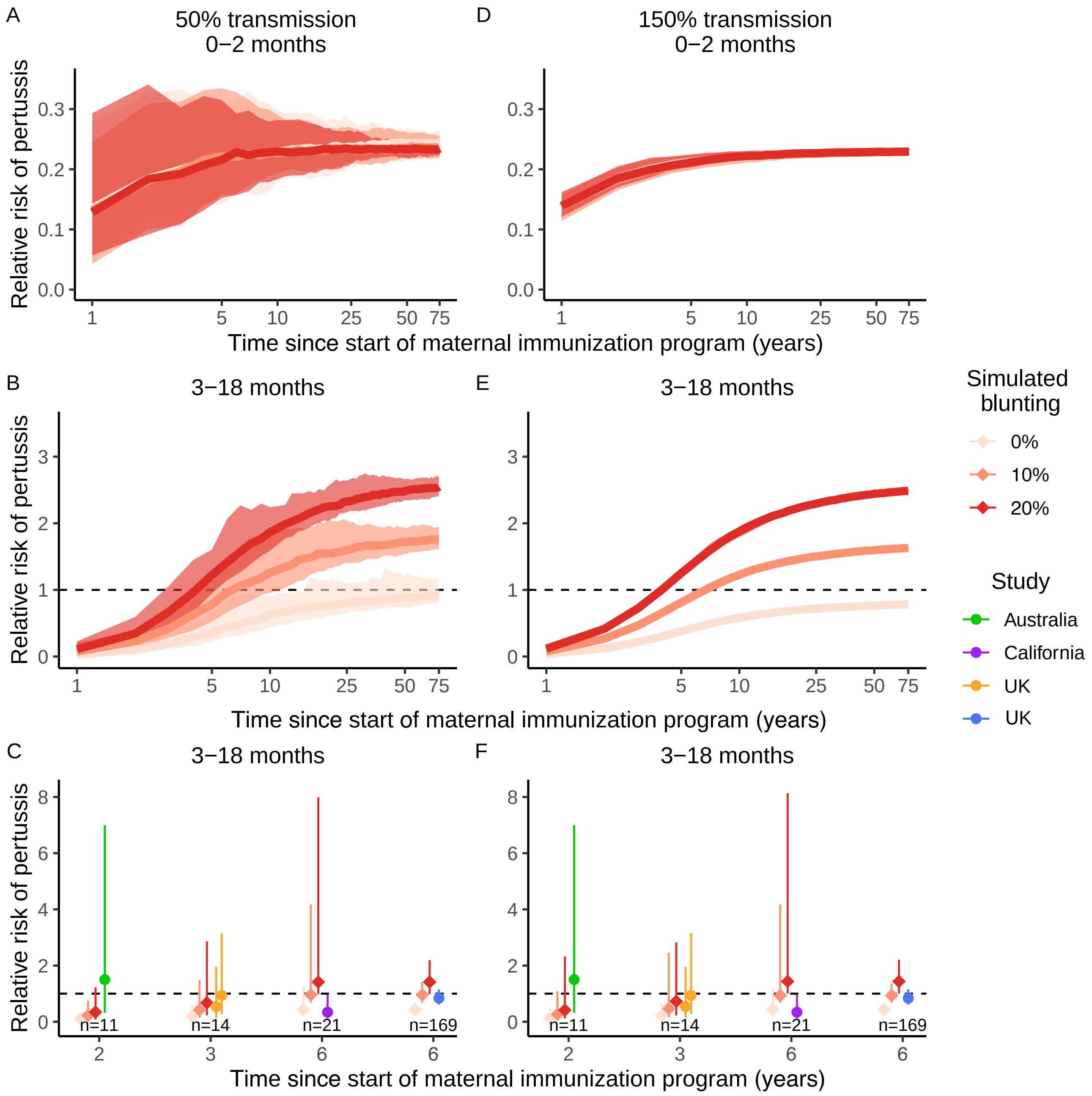


Figure S24: Sensitivity analysis 8: The RR of pertussis infection in infants from vaccinated relative to unvaccinated mothers in a population with the transmission in the 0 to 9-year-old being either low ($q_{1,\dots,11}$ $= 0.045$ in in panels A, B, C) or high ($q_{1,\dots,11} = 0.135$ in panels D, E, F). The baseline value in Fig. 5 is 0.09, which was the best-fitting value of the stochastic model on Massachusetts data [1]. All else is equal as in Fig. 5.

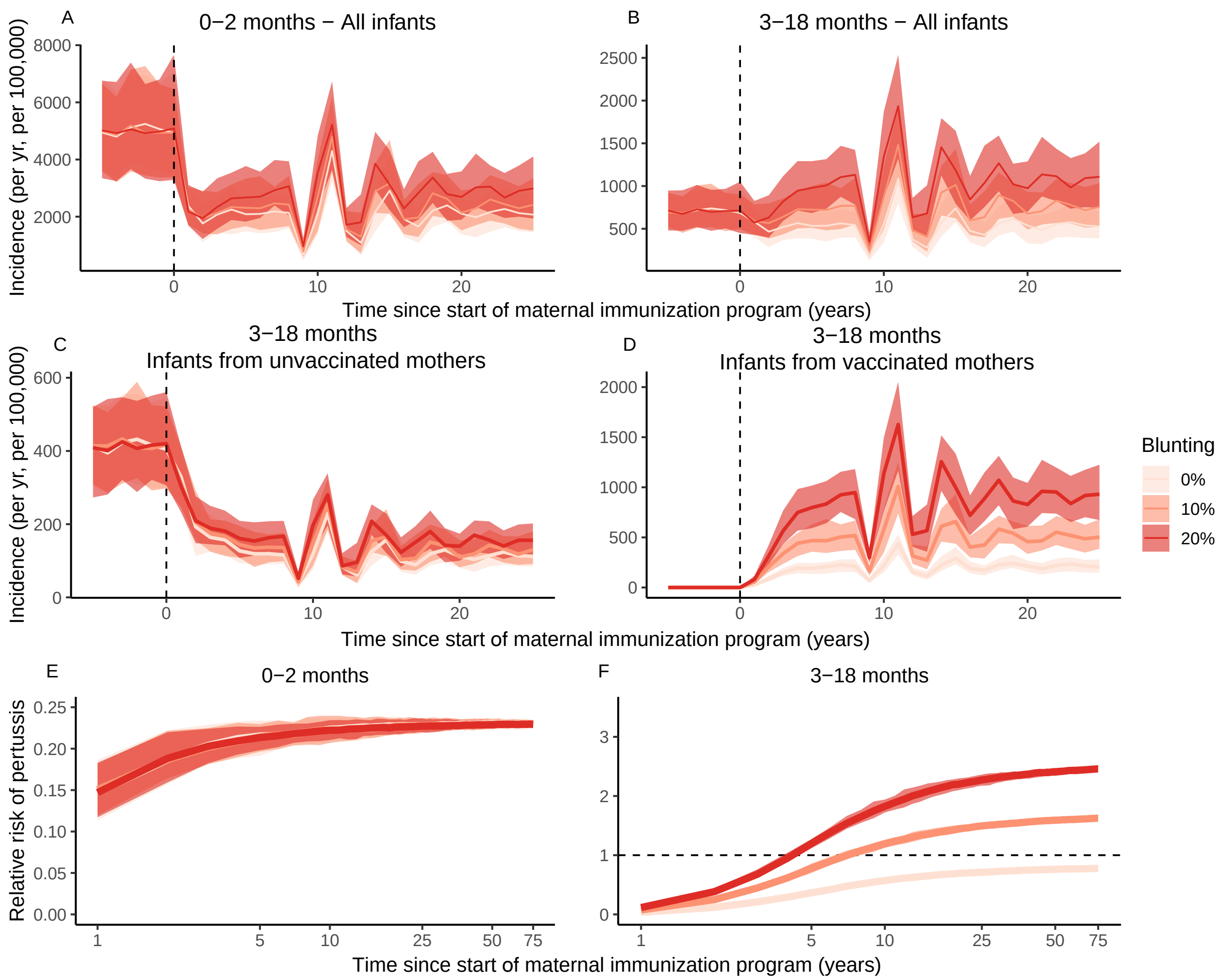


Figure S25: Sensitivity analysis 9: The consequences of 3 months of social distancing with a 20% reduction in transmission on pertussis incidence (A–D) and on the relative risk of pertussis following maternal immunization (E, F). All else is equal as in Fig. 4 and Fig. 5.

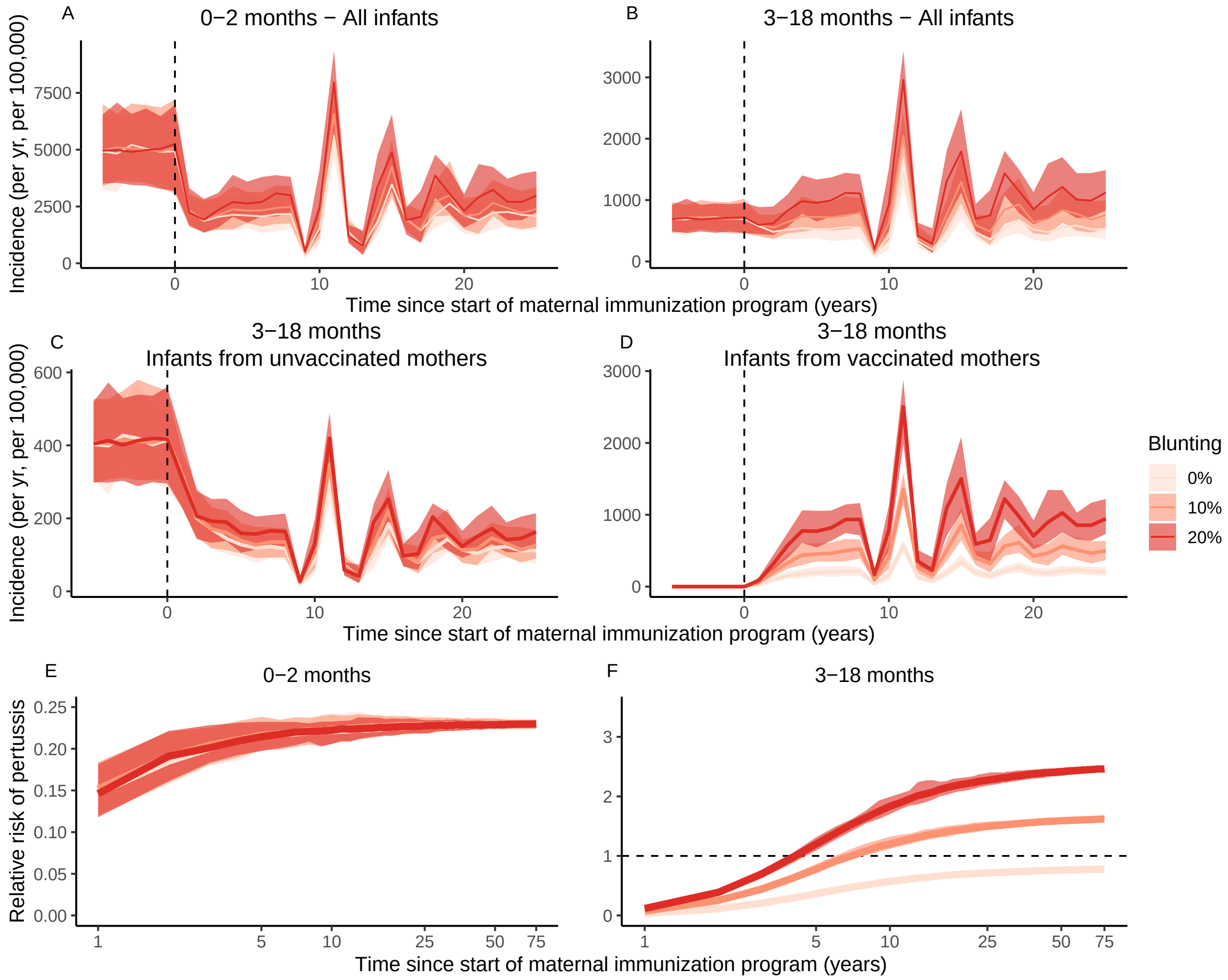


Figure S26: Sensitivity analysis 9: The consequences of 3 months of social distancing with a 50% reduction in transmission on pertussis incidence (A–D) and on the relative risk of pertussis following maternal immunization (E, F).All else is equal as in Fig. 4 and Fig. 5.

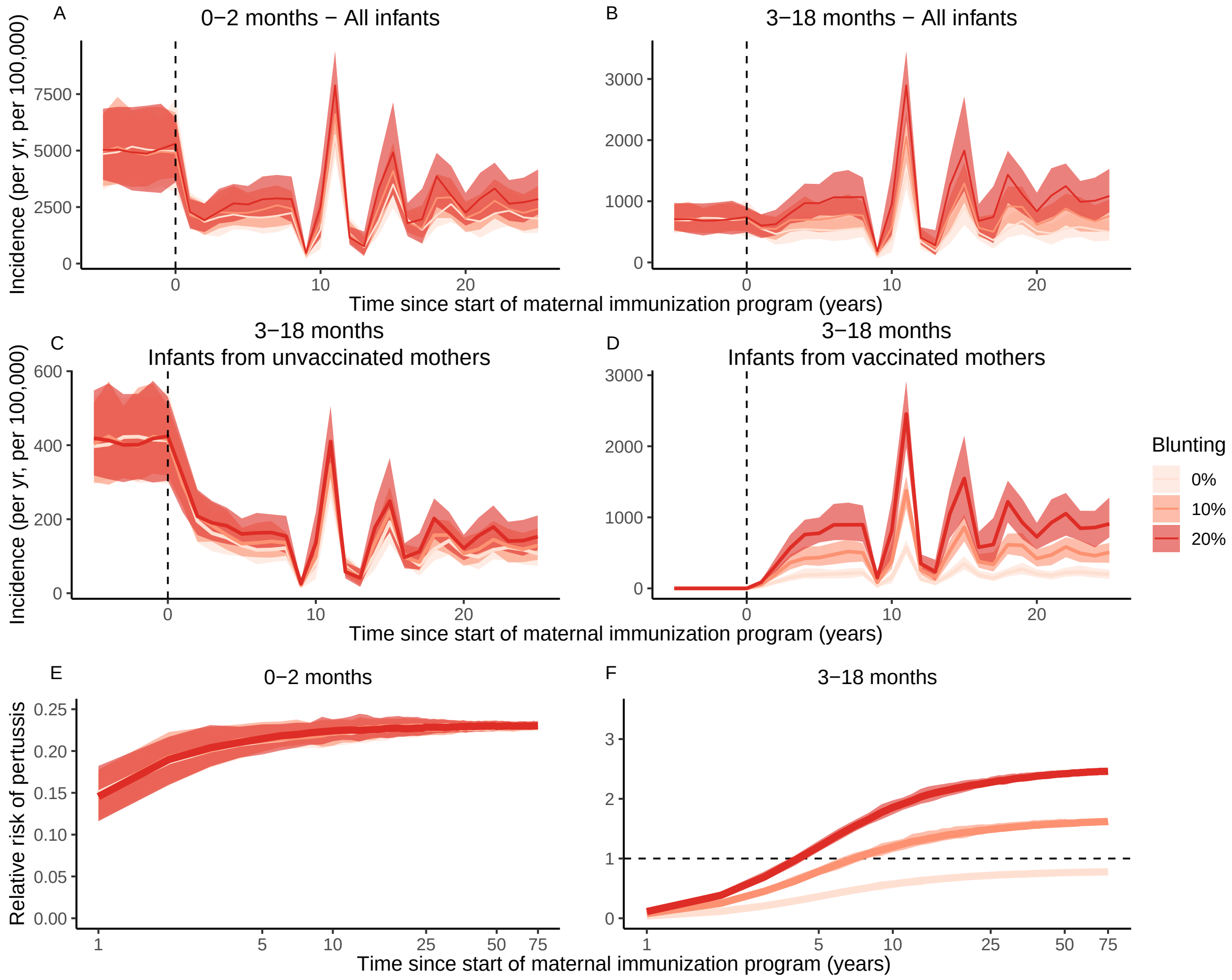


Figure S27: Sensitivity analysis 9: The consequences of 3 months of social distancing with a 70% reduction in transmission on pertussis incidence (A–D) and on the relative risk of pertussis following maternal immunization (E, F). All else is equal as in Fig. 4 and Fig. 5.

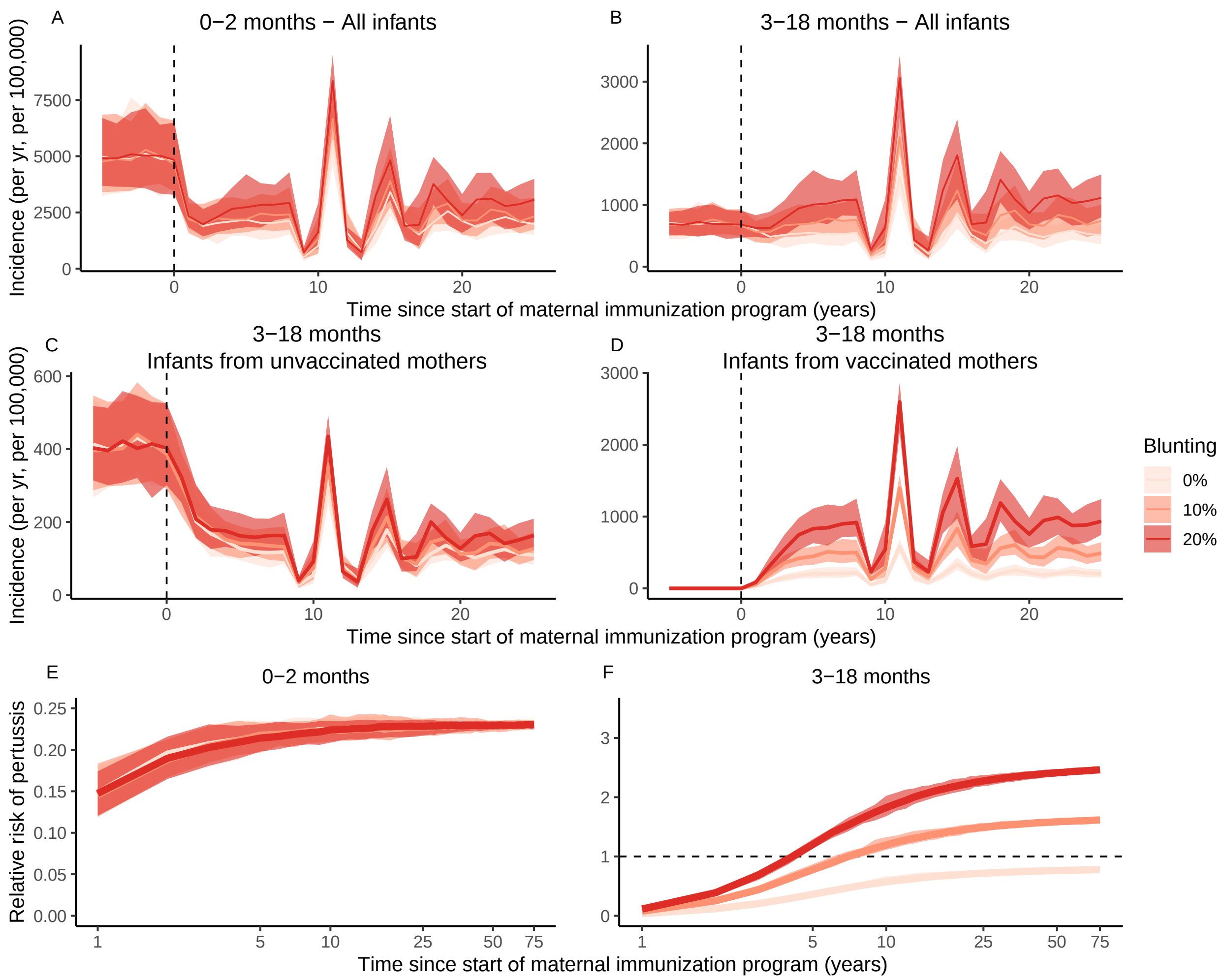


Figure S28: Sensitivity analysis 9: The consequences of 9 months of social distancing with a 20% reduction in transmission on pertussis incidence (A–D) and on the relative risk of pertussis following maternal immunization (E, F).All else is equal as in Fig. 4 and Fig. 5.

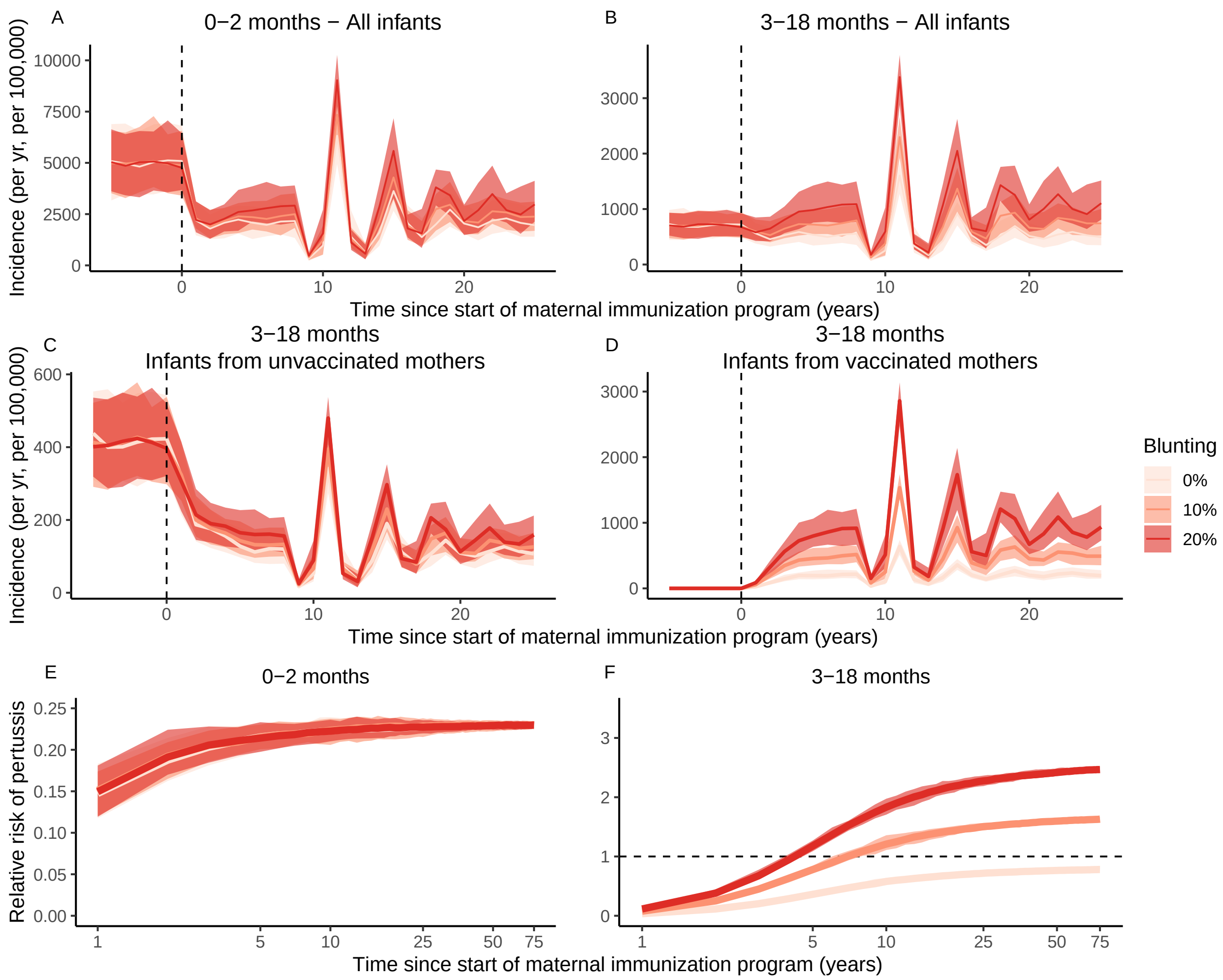


Figure S29: Sensitivity analysis 9: The consequences of 9 months of social distancing with a 50% reduction in transmission on pertussis incidence (A–D) and on the relative risk of pertussis following maternal immunization (E, F).All else is equal as in Fig. 4 and Fig. 5.

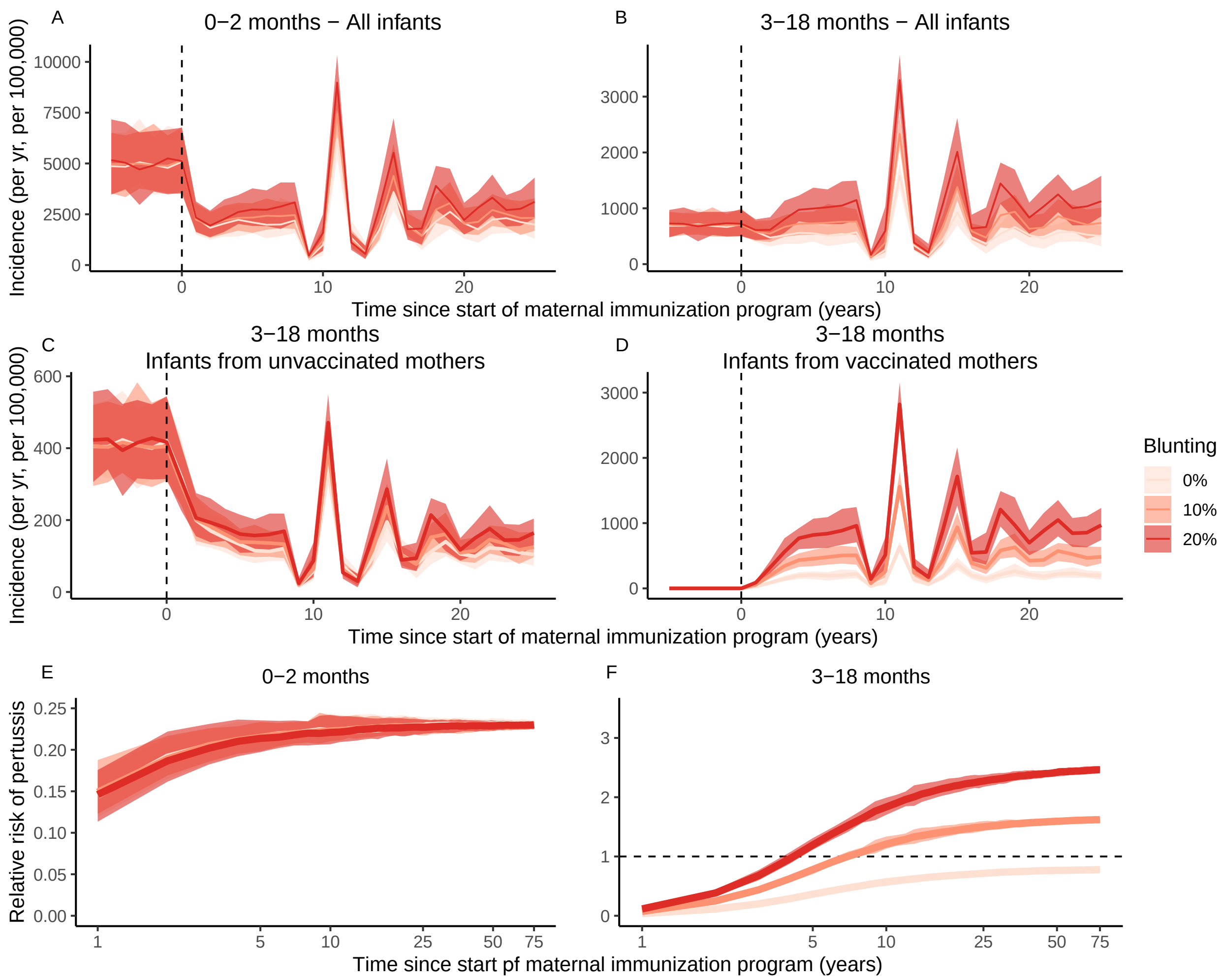


Figure S30: Sensitivity analysis 9: The consequences of 9 months of social distancing with a 70% reduction in transmission on pertussis incidence (A–D) and the relative risk of pertussis following maternal immunization (E, F).All else is equal as in Fig. 4 and Fig. 5.

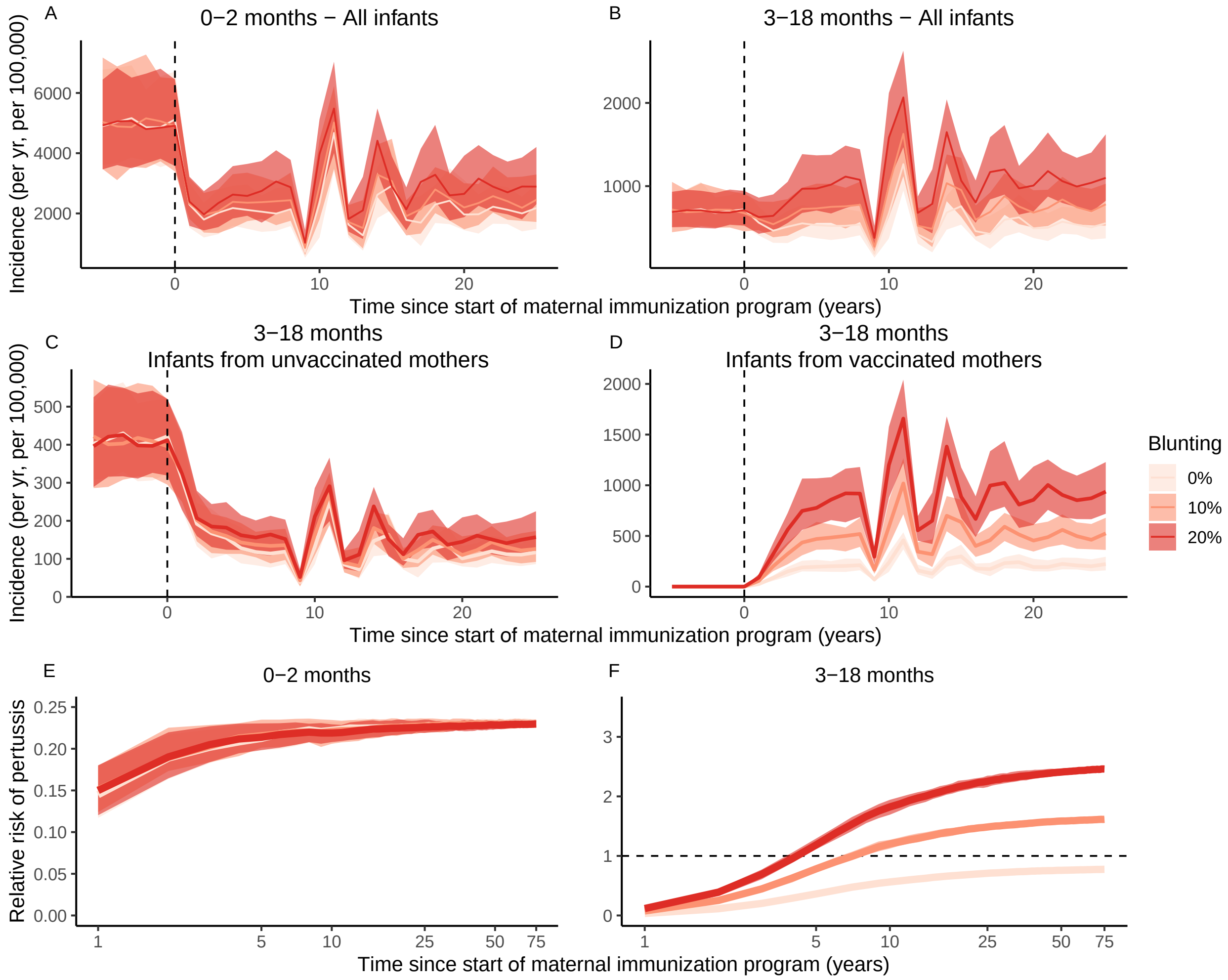


Figure S31: Sensitivity analysis 9: The consequences of 3 months of social distancing with a 20% reduction in transmission and a 20% reduction in immunization coverage on pertussis incidence (A–D) and the relative risk of pertussis following maternal immunization (E, F). All else is equal as in Fig. 4 and Fig. 5.

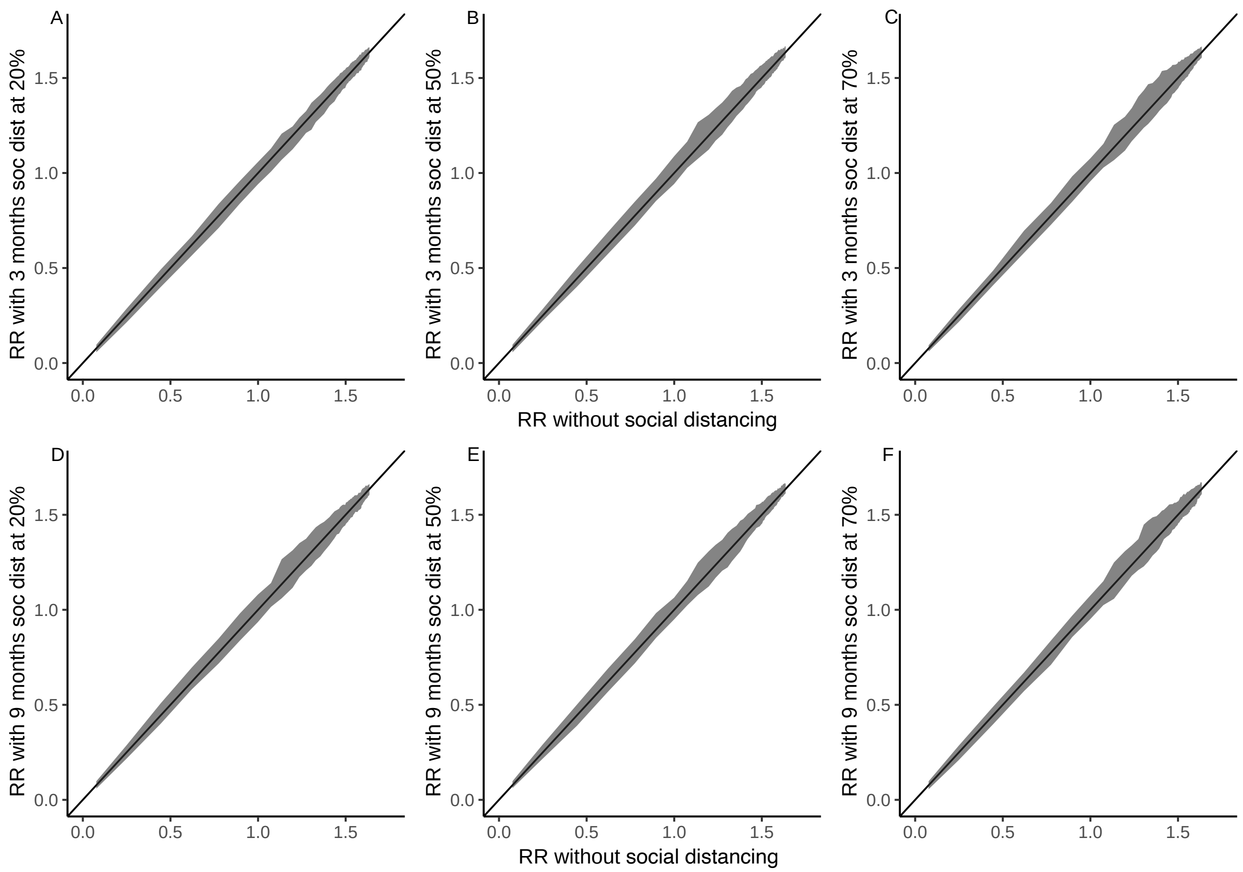


Figure S32: Sensitivity analysis 9: RR of pertussis in infants from vaccinated mothers relative to unvaccinated mothers without social distancing (on X-axis, i.e., the scenario in Fig. 4 and Fig. 5) and with various social distancing scenarios (soc dist on Y-axis). For illustration purposes we selected simulations with 10% blunting for both the X and Y axes. Variation around the first diagonal reflects stochasticity between simulations. Figures show results of 100 stochastic simulations.

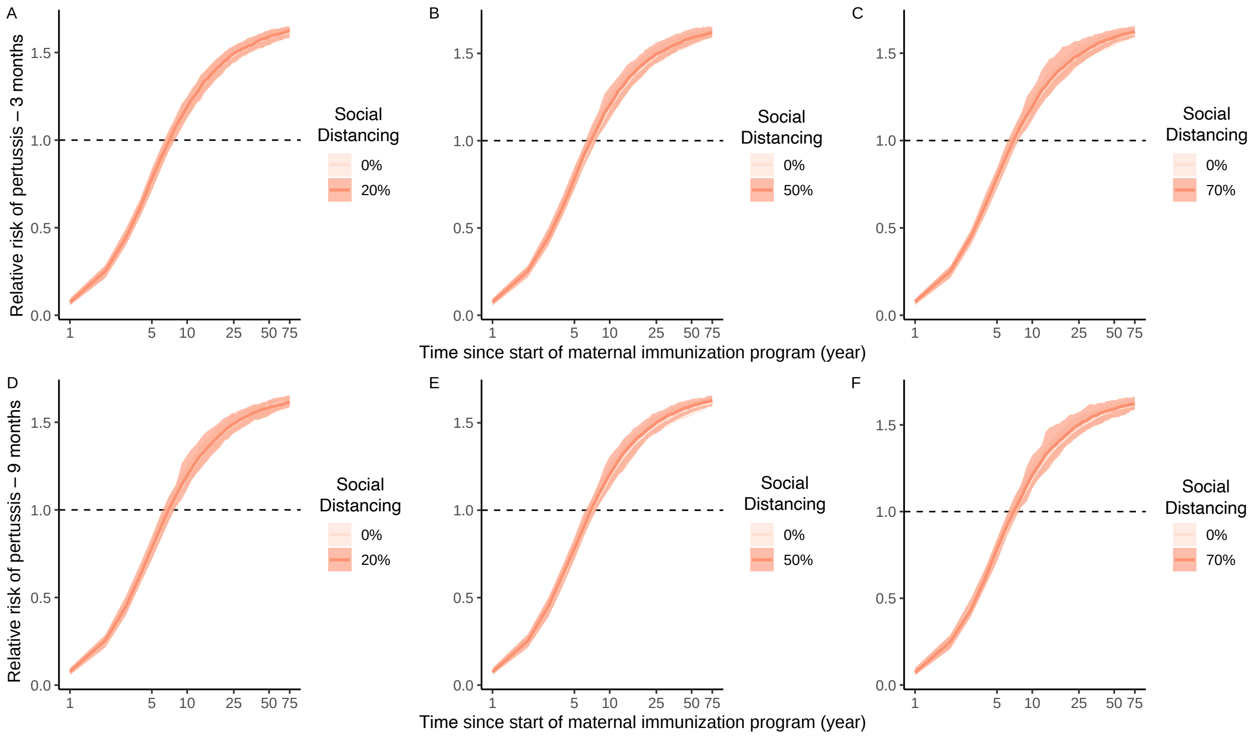


Figure S33: Sensitivity analysis 9: RR of pertussis in infants from vaccinated mothers relative to unvaccinated mothers without social distancing (i.e., the scenario in Fig. 4 and Fig. 5) and with various social distancing scenarios. For illustration purposes we selected simulations with 10% blunting for both with and without social distancing. Figures show the results of 100 stochastic simulations, with the solid line representing the median and the shaded areas indicating the 95% CI.

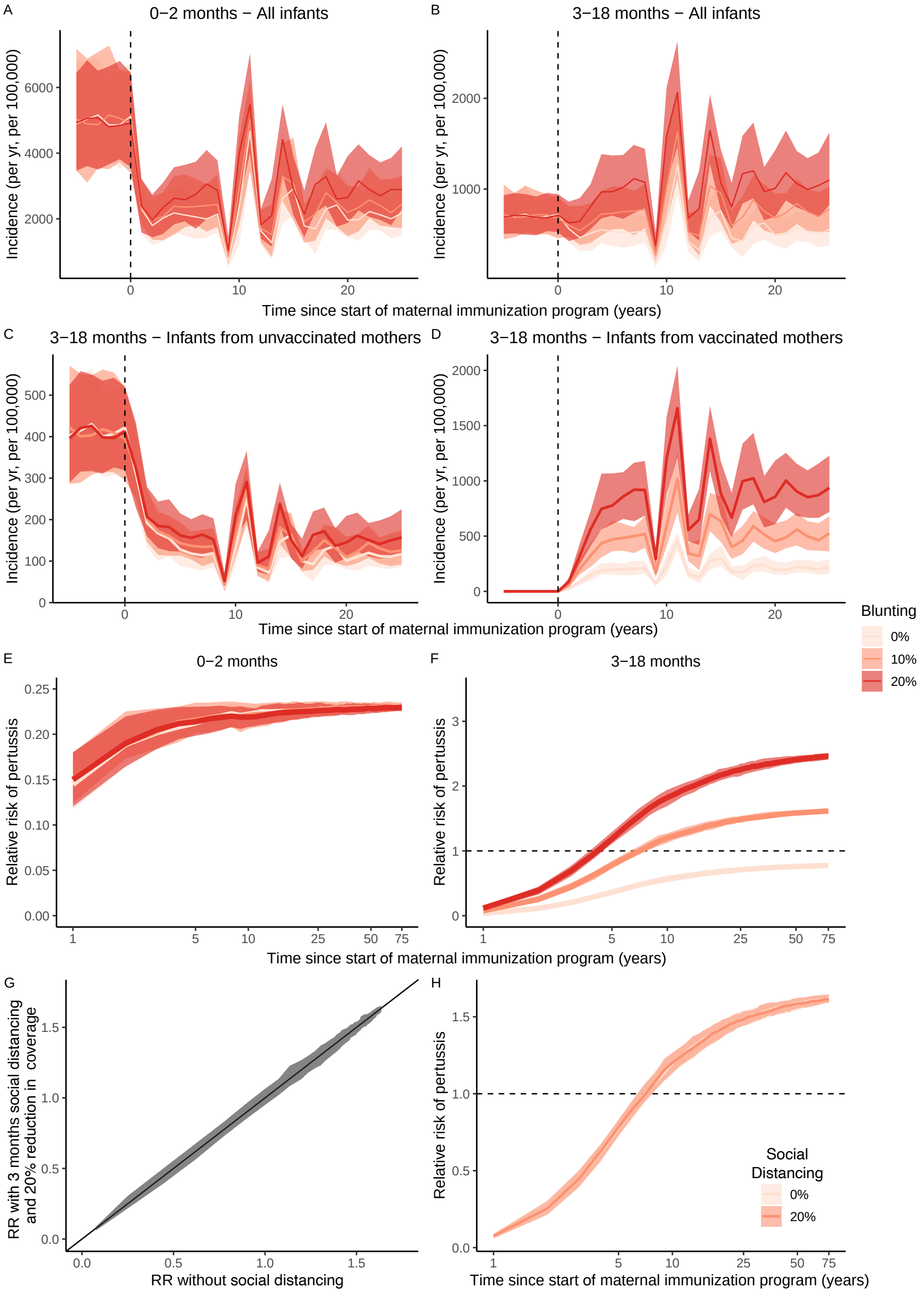


Figure S34: Sensitivity analysis 9: RR of pertussis in infants from vaccinated mothers relative to unvaccinated mothers with social distancing that has a 20% reduction in both transmission and coverage of all immunizations. Panels G and H compare this social distancing scenario relative to a scenario without social distancing (i.e., as in Fig. 4 and Fig. 5). In panels G and H, we selected, for illustration purposes, 10% blunting for both with and without social distancing. In panel (G), the variation around the first diagonal reflects stochasticity between simulations. Figures show the results of 100 stochastic simulations, with the solid line representing the median and the shaded areas indicating the 95% CI.

#### S3 Supplementary tables

| Study Ref | Publ Year | Country/ State | Start maternal vaccine year | Monitor years | Maternal coverage low [%] | Maternal coverage high [%] | Maternal coverage source |
|---|---|---|---|---|---|---|---|
| [6] | 2016 | UK | 10/2012 | 2013-2015 | CPRD: 50<br>ImmF: 50 | CPRD: 70<br>ImmF: 62 | Fig. 1 |
| [7] | 2017 | California | 10/2011 | 2010-2015 | 2010: 12<br>2012: 17 | 87 | Fig. 1 |
| [8] | 2021 | Australia | 06/2015 | 2015-2017 | 54 | 81 | Results paragr. 1,[15] |
| [9] | 2022 | UK | 10/2012 | 2012-2018 | 60 | 75 | Fig. 1 |
| **Study Ref** | **Primary series age [months]** | **Primary series cover [%]** | **Booster 1 age [months]** | **Booster 1 cover [%]** | **Booster 2 age [years]** | **Booster 2 cover [%]** | **Refs** |
| [6] | 2,3,4 | 90-95 | NA | NA | 3.5 | 85-97 | [16, 17] |
| [7] | 2,4,6 | 94 | 12-15 | 95 | 11-12 | 88 | [1, 18, 19] |
| [8] | 2,4,6 | 96 | 18 | 91 | 4 | 75-85 | [20, 21, 22] |
| [9] | 2,3,4 | 90-95 | NA | NA | 3.5 | 85-97 | [16, 17] |
| **Study Ref** | **Infant vaccine dose** | **Relative Risk** | | | **N cases** | **Source** | **Notes** |
| | | **95% lci** | **mean** | **95% uci** | | | |
| [6] | 1 | 0.09 | 0.19 | 0.43 | 32 | Table 5 | CPRD |
| [6] | 2 | 0.14 | 0.44 | 1.33 | 10 | Table 5 | CPRD |
| [6] | 3 | 0.15 | 0.54 | 1.96 | 14 | Table 5 | CPRD |
| [6] | 1 | 0.16 | 0.35 | 0.76 | 32 | Table 5 | ImmF |
| [6] | 2 | 0.25 | 0.80 | 2.56 | 10 | Table 5 | ImmF |
| [6] | 3 | 0.28 | 0.94 | 2.88 | 14 | Table 5 | ImmF |
| [7] | 1 | 0.06 | 0.19 | 0.58 | 28 | Table 5 | NA |
| [7] | 2 | 0.33 | 0.94 | 2.32 | 20 | Table 5 | NA |
| [7] | 3 | 0.12 | 0.34 | 0.96 | 21 | Table 5 | NA |
| [8] | 1 | 0.28 | 0.76 | 2.04 | 18 | Table 5 | NA |
| [8] | 2 | 0.32 | 1.50 | 7.00 | 11 | Table 5 | NA |
| [9] | 1 | 0.11 | 0.18 | 0.29 | 77 | Table 5 | NA |
| [9] | 2 | 0.25 | 0.52 | 1.11 | 28 | Table 5 | NA |
| [9] | 3 | 0.61 | 0.84 | 1.15 | 169 | Table 5 | NA |

Table S1: Data extracted and their references for Fig. 1 with their study design, vaccination schedules and coverages. Abbreviations: 95% lci = lower 95% confidence interval; 95% uci = upper 95% confidence interval

| State | Meaning |
|---|---|
| $V^{(M)}$ | Vaccinated from vaccinated mothers |
| $V^{(\bar{M})}$ | Vaccinated after failed maternal vaccination |
| $V$ | Vaccinated from unvaccinated mothers |
| $M$ | Protected by maternal Ab |
| $M^{(V)}$ | Protected by maternal Ab, after a primary vaccine failure |
| $S^{(\bar{M})}$ | Susceptible, after failed maternal vaccination |
| $S$ | Susceptible, from unvaccinated mothers |
| $E$ | Exposed |
| $I$ | Infected |
| $S^{(V)}$ | Susceptible, vaccinated from unvaccinated mothers |
| $S^{(V\bar{M})}$ | Susceptible, vaccinated from failed maternal vaccination) |
| $S^{(VM)}$ | Susceptible, vaccinated from successfully vaccinated mothers |
| $R$ | Recovered |

Table S2: List of state variables.

| Symbol | Meaning | Value (sensitivity analyses) | Source |
|---|---|---|---|
| | **Routine vaccination parameters** | | |
| $t_V$ | Start time of routine vaccination | 0 yrs | NA |
| $v_1$ | Primary infant immunization coverage | 90% (70%–80%) | [16, 19, 22] |
| $v_2$ | Booster infant immunization coverage | 90% (70%–80%) | [1, 17, 18, 20] |
| $\varepsilon$ | Vaccine effectiveness | 96% | [1] |
| $\alpha_V$ | Waning rate of vaccine protection | 0.011 $\text{yr}^{-1}$ | [1] |
| | **Maternal immunization parameters** | | |
| $t_M$ | Start time of maternal immunization | 100 yrs (60 yrs) | NA |
| $v_0$ | Maternal immunization coverage | 70% (50%–90%) | [6, 7, 8] |
| $\varepsilon_M$ | Effectiveness of maternal vaccination | 95% | [6, 7, 11, 23] |
| $\tau^{-1}$ | Average duration of maternally derived immunity | 8.7 months (4 mo–12 mo) | Calibrated to fit VE in [7, 11] |
| | **Blunting parameters** | | |
| $b_1$ | Blunting of vaccine effectiveness | 0–20% (40%–60%) | Calibrated to fit [6, 7, 8, 9] |
| $\bar{\varepsilon}$ | Blunted vaccine effectiveness $\varepsilon(1-b_1)$ | 76%–96% (46%) | Calibrated to fit [6, 7, 8, 9] |
| | **Transmission parameters** | | |
| $q_{1,\dots,11}$ | Susceptibility in 0–9 yr | 0.09 (0.045–0.135) | [1] |
| $q_{12,\dots,21}$ | Susceptibility in 10–19 yr | 0.05 (0.025–0.075) | [1] |
| $q_{22,\dots,76}$ | Susceptibility in $\geq 20$ yr | 0.008 (0.004–0.012) | [1] |
| $\sigma^{-1}$ | Average latent period | 8 days | [1] |
| $\gamma^{-1}$ | Average infectious period | 15 days | [1] |
| $C_{i,j}$ | Age-specific contact rates | POLYMOD | [12, 13] |
| $\iota$ | Prevalence of imported cases | 1 | [1] |
| | **Demographic parameters** | | |
| $\mu$ | Birth rate | 1/75 $\text{yr}^{-1}$ | [1] |
| $N$ | Total population size | $10^7$ | |
| $\Delta a_i$ | Span of age group $i$ | $\Delta a_1 = 2$ mo, $\Delta a_2 = 16$ mo, $\Delta a_3 = 6$ mo, $\Delta a_{i\geq 4} = 1$ yr | [1] |
| $N_i$ | Age-specific population sizes | $\frac{\Delta a_i}{75}N$ | [1] |
| $\delta_i$ | Aging rates | $1/\Delta a_i$ $\text{yr}^{-1}$ | [1] |

Table S3: List of model parameters for the main figures in the manuscript. Parameters values between brackets indicate the values tested in sensitivity analyses.
